# High-resolution portability of 245 polygenic scores when derived and applied in the same cohort

**DOI:** 10.1101/2021.02.05.21251061

**Authors:** Florian Privé, Hugues Aschard, Shai Carmi, Lasse Folkersen, Clive Hoggart, Paul F. O’Reilly, Bjarni J. Vilhjálmsson

## Abstract

The low portability of polygenic scores (PGS) across global populations is a major concern that must be addressed before PGS can be used for everyone in the clinic. Indeed, prediction accuracy has been shown to decay as a function of the genetic distance between the training and test cohorts. However, such cohorts differ not only in their genetic distance but also in their geographical distance and their data collection and assaying, conflating multiple factors. In this study, we examine the extent to which PGS are transferable between ancestries by deriving polygenic scores for 245 curated traits from the UK Biobank data and applying them in nine ancestry groups from the same cohort. By restricting both training and testing to the UK Biobank data, we reduce the risk of environmental and genotyping confounding from using different cohorts. We define the nine ancestry groups at a high-resolution, country-specific level, based on a simple, robust and effective method that we introduce here. We then apply two different predictive methods to derive polygenic scores for all 245 phenotypes, and show a systematic and dramatic reduction in portability of PGS trained in the inferred ancestral UK population and applied to the inferred ancestral Polish - Italian - Iranian - Indian - Chinese - Caribbean - Nigerian - Ashkenazi populations, respectively. These analyses, performed at a finer scale than the usual continental scale, demonstrate that prediction already drops off within European ancestries and reduces globally in proportion to PC distance, even when all individuals reside in the same country and are genotyped and phenotyped as part of the same cohort. Our study provides high-resolution and robust insights into the PGS portability problem.

## Introduction

Ever larger genetic data is becoming more readily available. This enables researchers to derive polygenic scores (PGS), which summarize an individual’s genetic component for a particular trait or disease by aggregating information from many genetic variants into a single score. In human genetics, polygenic scores are usually derived from summary statistics from a large meta-analysis of multiple Genome-Wide Association Studies (GWAS) and an ancestry-matched Linkage Disequilibrium (LD) reference panel (Choi *et al.* 2020). Polygenic scores can also be derived directly from individual-level data when available, i.e. from the genetic and phenotypic information of many individuals (De Los Campos *et al.* 2010). When using a single individual-level dataset with only moderate sample size, deriving polygenic scores usually results in poor prediction for most phenotypes, expect e.g. for autoimmune diseases with moderately large effects (Abraham *et al.* 2014; Privé *et al.* 2019). Fortunately, biobank datasets such as the UK Biobank now links genetic data for half a million individuals with phenotypic data for hundreds of traits and diseases (Bycroft *et al.* 2018). Thanks to the availability of these large datasets and to efficient methods recently developed to handle such data (Loh *et al.* 2018; Privé *et al.* 2019; Qian *et al.* 2020), individual-level data may be used to derive competitive PGS for hundreds of phenotypes.

A major concern about PGS is that they usually transfer poorly to other ancestries, e.g. a PGS derived from individuals of European ancestry is not likely to predict as well in individuals of African ancestry. Prediction in another ancestry has been shown to decay with genetic distance to the training population (Scutari *et al.* 2016; Wang *et al.* 2020) and with increasing proportion of admixture with a distant ancestry (Bitarello and Mathieson 2020; Cavazos and Witte 2020). This portability issue is suspected to be primarly due to differences in LD and allele frequencies between populations, and not so much about differences in effects and positions of causal variants (Shi *et al.* 2020; Wang *et al.* 2020; Cavazos and Witte 2020). Individual-level data from the UK Biobank offers an opportunity to further investigate this problem of PGS portability in a more controlled setting (Wang *et al.* 2020; Sinnott-Armstrong *et al.* 2021). Indeed, while the UKBB data contains genetic information for more than 450K British or European individuals, it also contains the same data for tens of thousands of individuals of non-British ancestry (Bycroft *et al.* 2018). Of particular interest, those individuals of diverse ancestries all live in the UK and had their genetic and phenotypic information derived in the same way as people of UK ancestry. This allows to circumvent potential confounding bias that might arise in comparative analyses from independent studies, and makes the UK Biobank data very well suited for comparing and evaluating predictive performance of derived PGS in diverse ancestries and across multiple phenotypes.

To investigate portability of PGS to other ancestries, we must first define groups of different ancestries from the data. Principal Component Analysis (PCA) has been widely used to correct for population structure in association studies and has been shown to mirror geography in Europe (Price *et al.* 2006; Novembre *et al*. 2008). Due to its popularity, many methods have been developed for efficiently performing PCA (Abraham *et al.* 2017; Privé *et al.* 2018, 2020a) as well as appropriately projecting samples onto a reference PCA space (Zhang *et al.* 2020a; Privé *et al.* 2020a), making it possible to perform these analyses for ever increasing datasets. Naturally, PCA has also been used for ancestry inference (Chen *et al.* 2013; Byun *et al.* 2017; Zhang *et al.* 2020a). However, among all studies where we have seen PCA used for ancestry inference, there does not seem to be a consensus on what is the most appropriate method for inferring ancestry using PCA. For example, there are divergences on which distance metric to use and the number of PCs to use to compute these distances. The ancestry of an individual can also be inferred based on other approaches, including the ADMIXTURE model, its various extensions, and haplotype-based methods (Alexander *et al*. 2009; Lawson *et al.* 2012; Raj *et al.* 2014; Frichot *et al.* 2014; Haller *et al.* 2017; Cheng *et al.* 2017; Jin *et al*. 2019; Cabreros and Storey 2019). However, we focus on PCA here because it is very fast.

In this study, we examine the extent to which PGS are transferable between ancestries by deriving 245 polygenic scores from the UK biobank data and applying them in nine ancestry groups from the same cohort. We first propose simple, robust and effective methods for global ancestry inference and grouping from PCA of genetic data, and use them to define nine ancestry groups in the UK Biobank data. We then apply a computationally efficient implementation of penalized regression (Privé *et al.* 2019) to derive PGS for 245 traits using the UK Biobank genetic and phenotypic data only. As an alternative method, we also run LDpred2-auto (Privé *et al.* 2020b), for which we directly derive the summary statistics from the individual-level data available. We show a dramatically low portability of PGS from UK ancestry to other ancestries. For example, on average, the phenotypic variance explained by the PGS is only 64.7% in India, 48.6% in China, and 18% in Nigeria compared to in individuals of UK ancestry. These results are presented at a finer scale than the usual continental level, which allows us to show that prediction already drops within Europe, e.g. for East and South Europe compared to UK. We find that this decay in variance explained by the PGS is roughly linear in the PC distance to the training population, and is remarkably consistent across most phenotypes and for both prediction methods applied. The few exceptions include traits such as hair color, tanning, and some blood measurements. We also explore using more than HapMap3 variants when fitting PGS, it proves useful when large effects are poorly tagged by HapMap3 variants, e.g. for lipoprotein(a), but not in the general case. We also explore the performance of PGS trained using a mixture of European and non-European ancestry samples, but do not observe any significant gain in prediction here.

## Results

### Overview of study

Here, we use the UK Biobank (UKBB) data only (Bycroft *et al*. 2018). We first infer nine ancestry groups in the UKBB. Then we use 391,124 individuals of UK ancestry to train polygenic scores (PGS) for 245 phenotypes (about half being diseases, see categories in figure S1) based on UKBB individual-level genotypes and phenotypes, and assess portability of these PGS in the remaining individuals of diverse ancestries (Table 1). As additional analyses, we also investigate using more variants than the HapMap3 variants used in the main analyses, and train models using a mixture of multiple ancestries. To derive PGS in this study, we use two different methods, penalized regression and LDpred2-auto, and finally compare them.

**Table 1:** In total, 439,378 unrelated individuals are used here. Main analyses use UK1 + UK2 (391,124 individuals) as training set and the other groups as test sets. Secondary analyses involve multiple ancestry training and keep only the UK3 and Caribbean groups as test sets; UK2 is removed from the training so that sample size from training 2 is the same as training 1 (391,124 individuals).

| Set | UK1 | UK2 | UK3 | Poland | Italy | Iran | India | China | Caribbean | Nigeria | Ashkenazi |
| --- | --- | --- | --- | --- | --- | --- | --- | --- | --- | --- | --- |
| Training 1 | 367,063 | 24,061 |  |  |  |  |  |  |  |  |  |
| Test 1 |  |  | 20,000 | 4136 | 6660 | 1200 | 6331 | 1810 | 2484 | 3924 | 1709 |
| Training 2 | 367,063 |  |  | 4136 | 6660 | 1200 | 6331 | 1810 |  | 3924 |  |
| Test 2 |  |  | 20,000 |  |  |  |  |  | 2484 |  |  |

### Ancestry grouping

We investigate various approaches to classify individuals in ancestry groups based on Principal Component Analysis (PCA) of genome-wide genotype data. Detailed results can be found in the corresponding Supplementary Note; we recall main results here. First, we show that (squared) Euclidean distances in the PCA space of genetic data are approximately proportional to *F*_*ST*_ between populations, and we therefore recommend using this simple distance. At the same time, we provide evidence that using only 2 PCs, or even 4 PCs, is not enough to distinguish between some less-distant populations, and recommend using all PCs visually capturing some population structure. Then, we use this PCA-based distance to infer ancestry in the UK Biobank and the POPRES datasets. We propose two solutions to do so, either relying on projection of PCs to reference populations such as the 1000 Genomes Project, or by directly using internal data only. We show that these solutions are simple, robust and effective methods for inferring global ancestry and for grouping genetically homogeneous individuals.

Here, we first use the second solution presented in the Supplementary Note, relying on PCs computed within the UK Biobank and individual information on the countries of birth, for inferring the first eight ancestry groups presented in table 1. Then, for inferring the “Ashkenazi Jewish” ancestry group, we use the first solution, projecting UKBB individuals onto the PCA space of a reference dataset composed of many Jewish and non-Jewish individuals (Behar *et al.* 2013). We identify a ninth group of 1709 unrelated individuals, which is entirely disjunct from the other eight groups previously defined (Methods). The nine ancestry groups inferred here are represented in the UKBB PCA space in figure 1.

**Figure 1:**
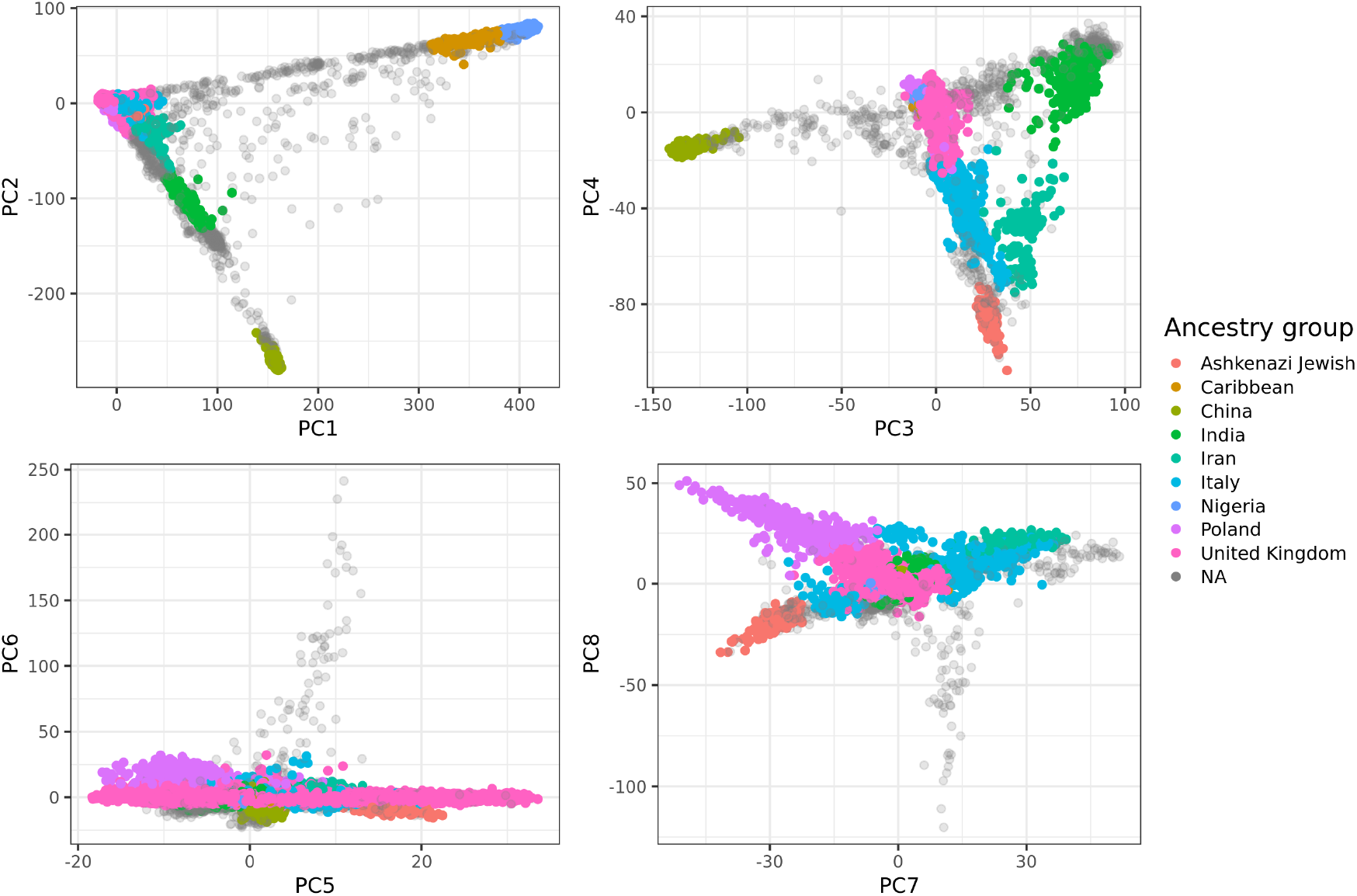
The first eight PC scores of the UK Biobank (Field 22009) colored by the homogeneous ancestry group we infer for these individuals. Only 50K random individuals are represented.

### Portability of polygenic scores to other ancestries

Figure 2 presents the results when fitting penalized regression using a training set composed of UK individuals only and testing in nine different ancestry groups. Averaged over 245 phenotypes, compared to prediction performance in individuals of UK ancestry, relative predictive ability in terms of partial-*r*^2^ (Methods) is 93.8% in Poland, 85.6% in Italy, 72.2% in Iran, 64.7% in India, 48.6% in China, 25.2% in the Caribbean, 18% in Nigeria, and 85.7% for the Ashkenazi group. As a follow-up analysis to ensure that this drop in performance in other ancestries is not due to imputation, we perform the same analysis for 83 of the continuous phenotypes using high-quality genotyped variants only (Methods) instead of the (mostly imputed) HapMap3 variants; results are highly consistent (Figure S2). These results are also very similar when using LDpred2-auto instead of penalized regression for training predictive models for all phenotypes (Figure S3). A few phenotypes deviate from this global trend, e.g. prediction of bilirubin concentration ranges between 0.537 and 0.619 (partial-*r*) for all ancestries except for China, for which it is 0.415 (95% CI: 0.374 - 0.453, see Methods). In contrast, e.g. for hair and skin color, partial correlations decrease quickly and are not significantly different from 0 for both China and Nigeria, while of 0.420 (95% CI: 0.409 - 0.432) for “darker hair” in the UK ancestry group (Figure 2). Overall, relative predictive performance decreases approximately linearly with PC distance to the UK (Figure 3). A similar pattern is observed when computing PCA based on more balanced ancestry groups, as recommended in Privé *et al.* (2020a) (Figure S4).

**Figure 2:**
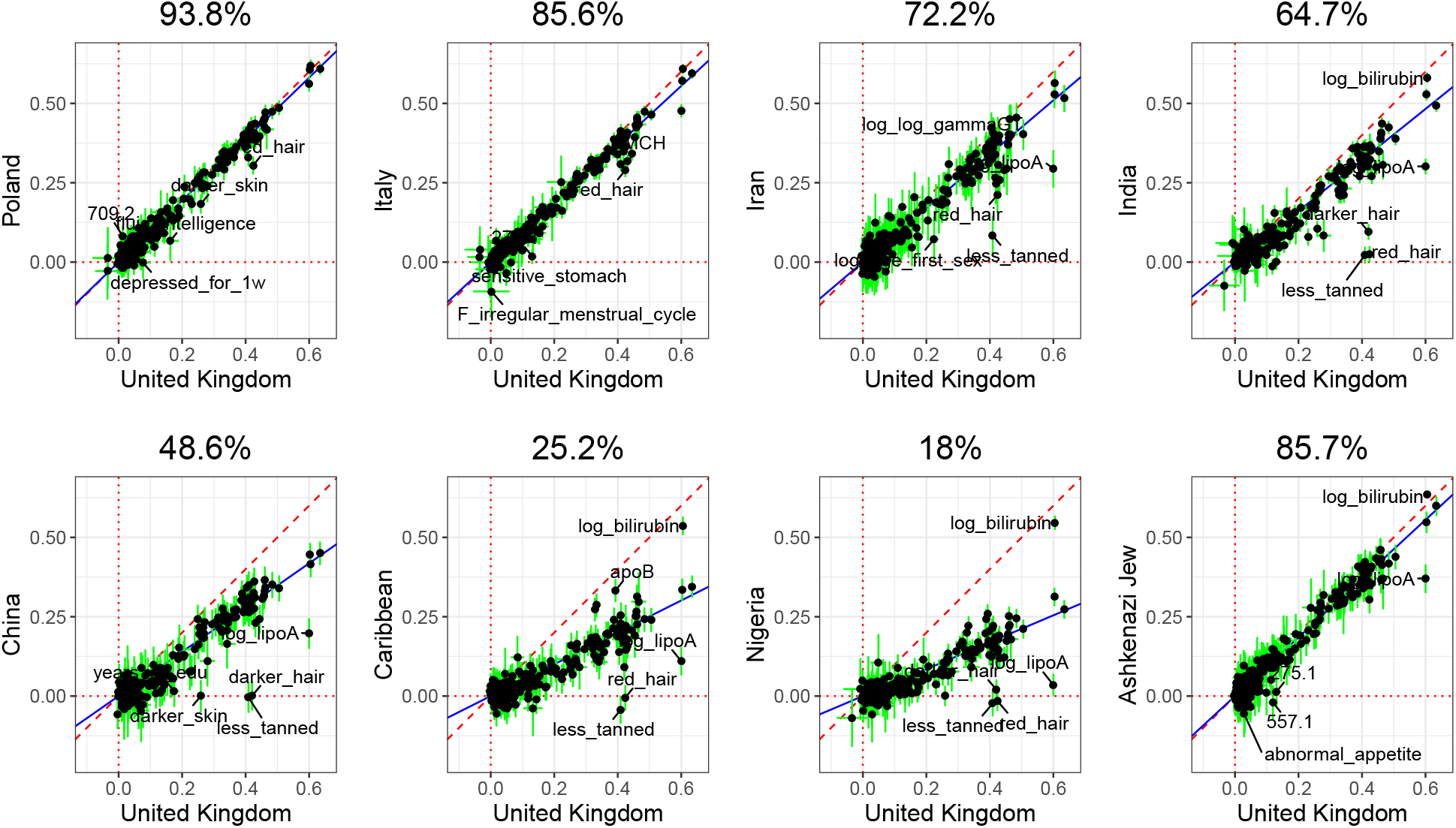
Partial correlation (and 95% CI) in the UK test set versus in a test set from another ancestry group (each panel). Each point represents a phenotype and training has been performed with penalized regression on UK individuals (training 1 in table 1) and HapMap3 variants. The slope (in blue) is computed using Deming regression accounting for standard errors in both x and y, fixing the intercept at 0. The square of this slope is provided above each plot, which we report as the relative predictive performance compared to testing in UK.

**Figure 3:**
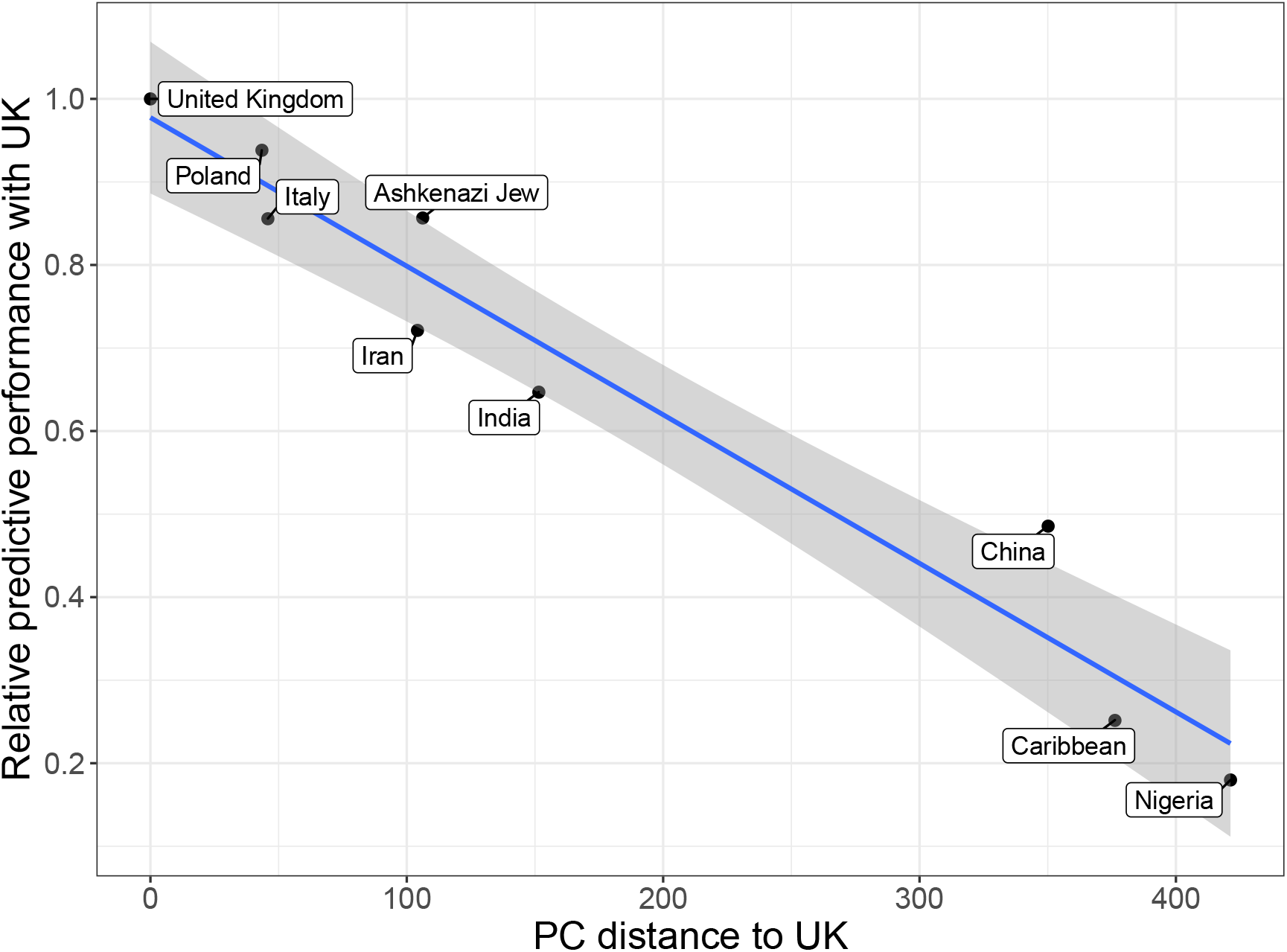
Relative predictive performance compared to the UK (ratio of variance explained in one group compared to in the UK group) versus PC distance from the UK. PC distances are computed using Euclidean distance between geometric medians of the first 16 reported PC scores (Field 22009) of each ancestry group. Relative performance values are the ones reported in figure 2. The slope and standard errors are computed internally by function `geom_smooth(method = “lm”)` of R package ggplot2.

### Using more than HapMap3 variants?

We investigate some of the outlier phenotypes in figure 2, especially the ones from blood biochemistry which have some variants with large effects. We hypothesize that using a denser set of variants could improve tagging of the causal variants with large effect sizes, resulting in an improved prediction in all ancestries. We focus on “total bilirubin”, “lipoprotein(a)” (lipoA) and “apolipoprotein B” (apoB). We perform a localized GWAS which includes all variants around the most significant variant (hereinafter denoted as “top hit”) from the GWAS in the training set 1 (UK individuals and HapMap3 variants only) in each of the first eight ancestry groups defined here. More precisely, we include all variants with an imputation INFO score larger than 0.3 and within a window of 500Kb from the HapMap3 top hit in the UK; there are approximately 30K such variants for all three phenotypes. For bilirubin, the overall top hit is a HapMap3 variant and explains around

30% of the phenotypic variance (Figure S6). Effects from the three top hits are fairly consistent within all ancestry groups (Figure S7) explaining why genetic prediction is highly consistent in all ancestries, except for China (Figure 2), for which these variants are rarer. For lipoA, results are very different across ancestries; HapMap3 variants are far from being the top hits for the UK individuals, where the top HapMap3 variant explains 5% of phenotypic variance compared to 29% for the (non-HapMap3) top hit (Figure 4). Note that this top hit is more than 200Kb away from the HapMap3 top hit from UK. Moreover, the 3 top hits for lipoA do not have very consistent effect sizes across ancestries (Figure S8). Finally, for apoB, effects from the three top hits, which are not part of HapMap3 variants, are fairly consistent across ancestries and explain up to 8.5% of the phenotypic variance (Figures S9 & S10).

**Figure 4:**
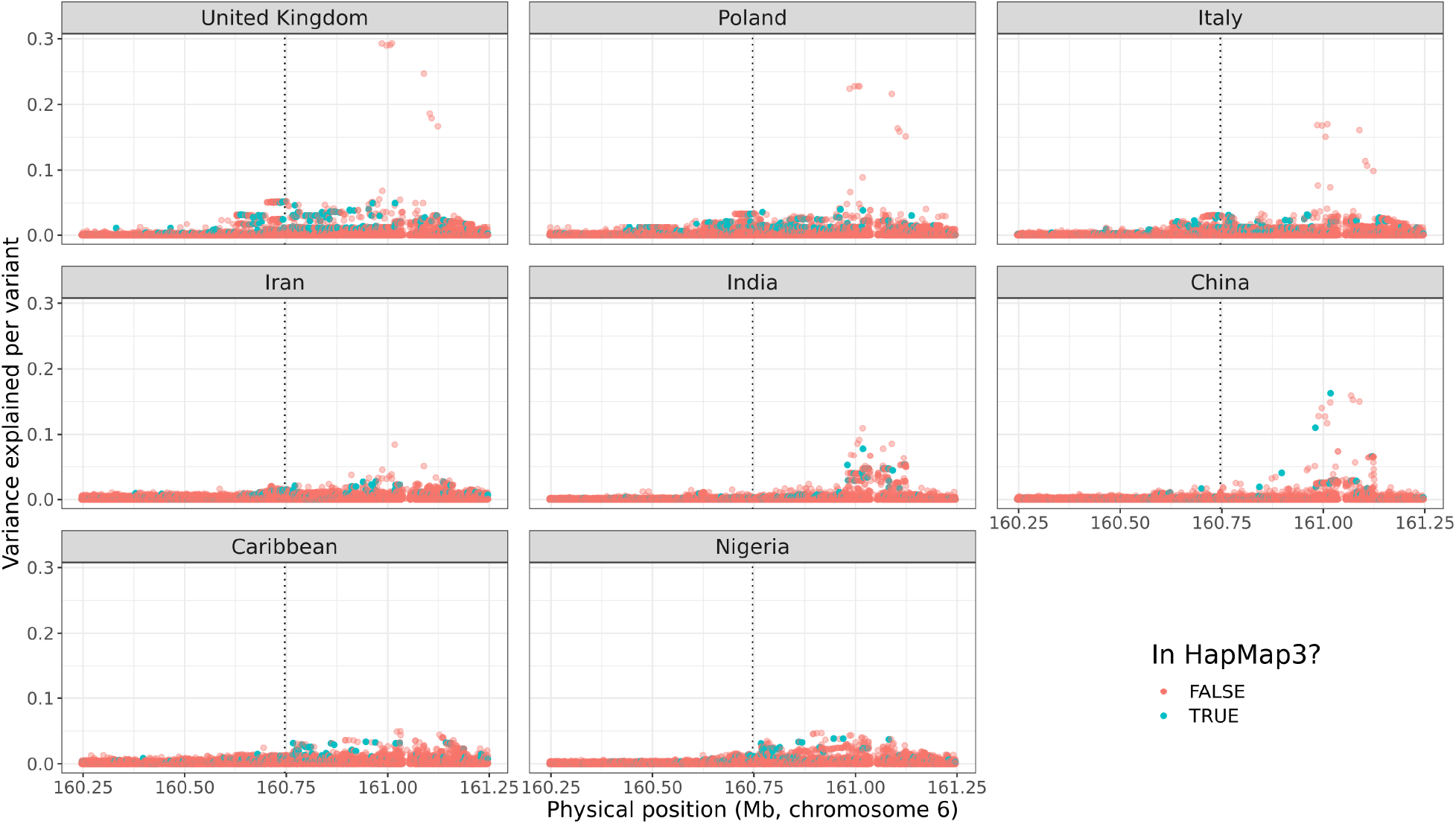
Zoomed Manhattan plot for lipoprotein(a) concentration. The phenotypic variance explained per variant is computed as *r*^2^ = *t*^2^*/*(*n* + *t*^2^), where *t* is the t-score from GWAS and *n* is the degrees of freedom (the sample size minus the number of variables in the model, i.e. the covariates used in the GWAS, the intercept and the variant). The GWAS includes all variants with an imputation INFO score larger than 0.3 and within a 500Kb radius around the top hit from the GWAS performed in the UK training set and on the HapMap3 variants, represented by a vertical dotted line.

We then investigate if the use of a larger set of variants than the HapMap3 set is beneficial; we use more than 8M common variants (Methods) and apply LDpred2-auto after restricting to the 1M most significant variants and applying winner’s curse correction (Methods). Except for lipoA for which we get a large improvement in predictive accuracy compared to using HapMap3 variants only, it is not beneficial for the other seven phenotypes analyzed here (Figure 5). Remarkably, while the partial correlation for lipoA is about 75% in the UK test set when using this prioritized set of variants, it is still not different from 0 when applied to the Nigeria group. For height and BMI, estimated SNP heritability is reduced when using this set of most significant variants only, and all these variants are estimated to be causal, i.e. estimate of *p* is 1

**Figure 5:**
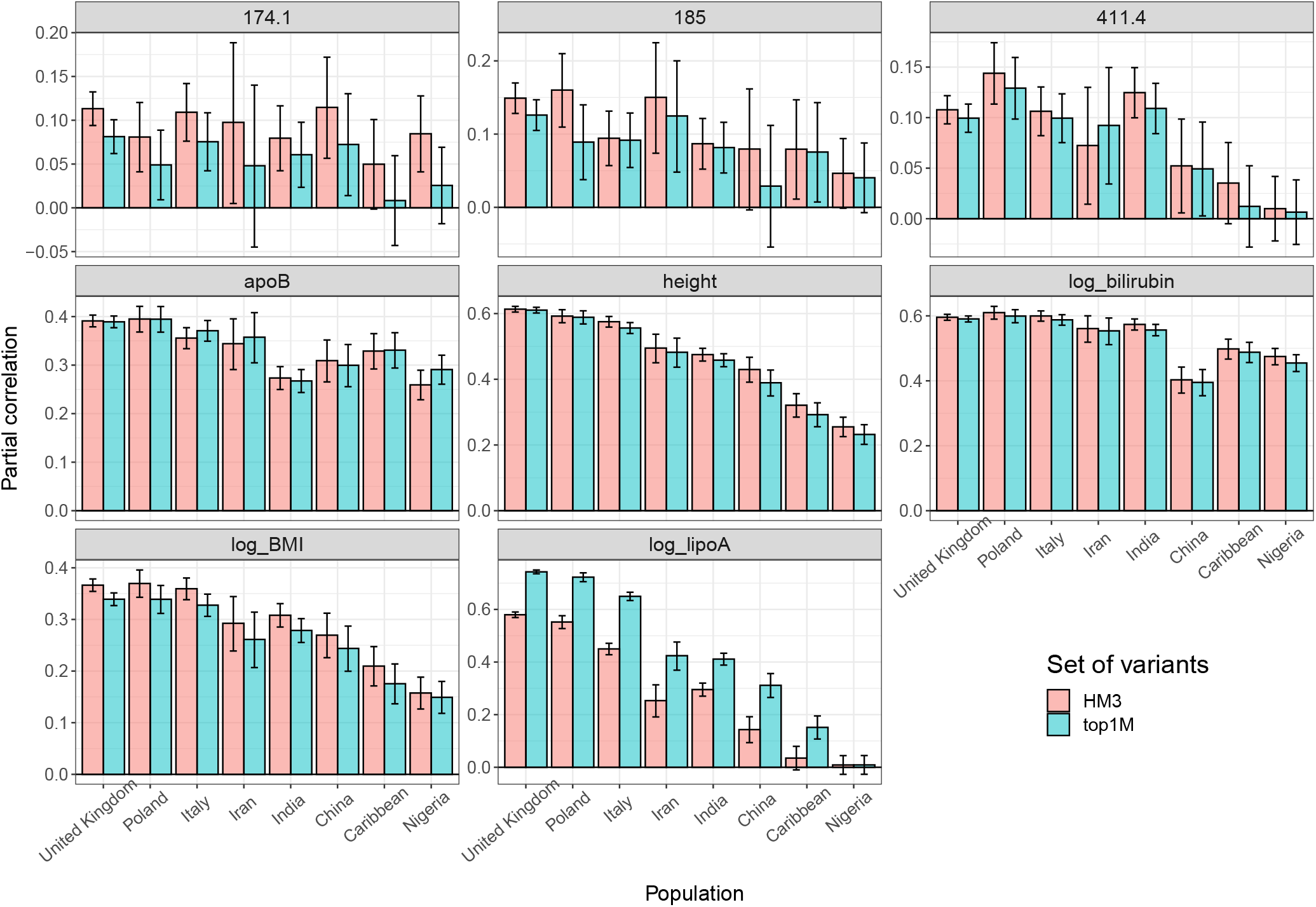
Predictive performance with LDpred2-auto for 8 phenotypes (each panel), when using either HapMap3 variants (HM3) or the 1M most significant variants (top1M) out of more than 8M common variants (see Methods). Phecode 174.1: breast cancer; 185: prostate cancer; 411.4: coronary artery disease.

(Table S1). As height and BMI are very polygenic traits (*p* is estimated to be ∼2% and ∼4% respectively when using HapMap3 variants), contribution from less significant causal variants is missed due to this thresholding selection. For the three binary phenotypes, breast cancer (phecode: 174.1), prostate cancer (185) and coronary artery disease (411.4), although heritability estimates are larger when using this set of prioritized variants (Table S1), predictive accuracy does not improve compared to when using HapMap3 variants (Figure 5).

### Training with a mixture of ancestries

Here we use all ancestry groups except for the Caribbean and Ashkenazi for training; we remove the same number of UK individuals to keep the same training sample size as before (training 2 in table 1). We recall that Caribbean individuals are mostly admixed between African, European and Native American ancestries (Moreno-Estrada *et al.* 2013), which are almost all represented here in the training set 2. In figure S11, we investigate nine phenotypes of interest, either because they are highly studied diseases or are outliers in figure 2: breast cancer (phecode: 174.1), prostate cancer (186), type-2 diabetes (250.2), hypertension (401), coronary artery disease (411.4), skin tone, total bilirubin concentration, lipoprotein(a) concentration, and years of education. We predict in the test sets from the UK and the Caribbean (test set 2); overall, the predictive performance is highly similar when using this multi-ancestry training compared to when using only UK individuals, in both the UK and the Caribbean target samples. Prediction is only improved for lipoprotein(a) concentration when the mixed ancestry training data is used in application to the Caribbean target data (Figure S11).

### Comparison of predictive models

Penalized regression and LDpred2-auto provides approximately similar predictive performance across all traits and ancestries considered here (Figure S12); there are only four pairs of phenotype-ancestry (out of nearly 2000 pairs) for which 95% CIs for partial-*r* from penalized regression and LDpred2 are not overlapping: “615: Endometriosis” in China with 0.065 (0.0074 - 0.122) vs −0.051 (−0.108 - 0.0068); “hard falling asleep” in UK with −0.0349 (−0.742 - 0.0045) vs 0.071 (0.031 - 0.110); height in UK with 0.634 (0.626 - 0.643) vs 0.613 (0.605 - 0.622); log-bilirubin in Nigeria with 0.546 (0.523 - 0.569) vs 0.475 (0.449 - 0.500). For prediction in UK ancestry, penalized regression tends to provide better predictive performance than LDpred2 for phenotypes for which partial-*r* > 0.3, and LDpred2 tends to outperform penalized regression for phenotypes harder to predict (Figure S12).

Both methods allow for fitting sparse effects, i.e. some resulting effects are exactly 0. Sparse models may be beneficial because they may be more easily interpreted and implemented. The sparse option in LDpred2-auto provides similar performance as LDpred2-auto without this option (Figure S13). Sparsity of resulting effects follows a very different pattern for penalized regression compared to LDpred2-auto-sparse. Indeed, penalized regression tends not to include variants if it is uncertain that they have a non-zero effect, i.e. when effects are very small and prediction is difficult (Figure S14). In contrast, LDpred2-auto-sparse tends not to discard variants, only when *h*^2^ is large enough it sets lots of effects to 0 if *p* is small (Figure S15). Finally, running each penalized regression model takes between a few minutes and a few days depending on the number of non-zero effects in the resulting model (Figure S16). In contrast, LDpred2-auto should take the same computation time for all phenotypes; it completed under seven hours for most phenotypes (Figure S17).

## Discussion

In this paper, we have shown a poor portability of PGS to other populations, in agreement with what has been previously reported. Indeed, compared to three previous studies (Martin *et al.* 2019; Duncan *et al*. 2019; Wang *et al.* 2020), we show a relative predictive performance compared to Europeans of ∼18% for Africans (vs. 22%, 42% and 24%), ∼49% for East Asians (vs. 50%, 95% and 64%) and ∼65% for South Asians (vs. 60%, 62.5% and 72%). Our results provide a significant addition to the current literature in many ways. First, we show that the portability issue remains strong even when PGS are derived and applied in the same cohort. Second, the presented results are averaged over 245 phenotypes, which is much more than what has been typically used, and should capture a broad range of the phenotypic spectrum. Third, we provide this result at a finer scale than the usual continental level by proposing a simple, robust and effective method for grouping UKBB individuals in nine ancestry groups. This allows us to show e.g. that predictive performance already decreases within Europe with only ∼94% for East Europe and ∼86% for South Europe of the performance reached within UK.

We showcase two methods for deriving polygenic scores when large individual-level data is available. Although LDpred2-auto is a method based on summary statistics, it provides good predictive performance compared to penalized regression, when applied to individual-level data. Moreover, portability results shown here are similar when using either the individual-level penalized regression or the summary statistics based LDpred2 method. Fitting of penalized models is relatively fast when using 1M HapMap3 variants. We have also tried fitting penalized regression using 8M variants (>3TB of data); this was possible but took several days for the phenotypes we tried, so we have not investigated this further. To the best of our knowledge, the implementation we use is the most efficient penalized regression implementation currently available. Recently, Qian *et al.* (2020) proposed snpnet, a new R package for fitting penalized regressions on large individual-level genetic data, but we have found it to be much less efficient than R package bigstatsr on UKBB data (Supplementary Note). As for LDpred2, it currently cannot be run using 8M variants, but we show how to use a subset of 1M prioritized variants out of these 8M. Using this new set of variants provides a large improvement in predicting lipoprotein(a) concentration (lipoA), but not for the other seven phenotypes studied in this analysis. This improvement for lipoA is not surprising given that the top HapMap3 variant explains 5% of phenotypic variance compared to 29% for the (non-HapMap3) top hit (Figure 4).

Here we only use the UK Biobank data to fit polygenic scores. We do not use external information such as functional annotations; those could be used to improve the heritability model assumed by predictive methods in order to improve predictive performance (Zhang *et al.* 2020b). Moreover, we do not use external summary statistics. Nevertheless, Albiñana *et al.* (2020) have shown that an efficient strategy to improve predictive ability of polygenic scores consists in combining two different polygenic scores, one derived using external summary statistics, and another one derived using internal individual-level data. Therefore, the polygenic scores derived here could be combined with polygenic scores derived using external summary statistics; we will release these PGS publicly and share them in databases such as the PGS Catalog and the Cancer-PRSweb (Fritsche *et al.* 2020; Lambert *et al.* 2020).

## Materials and Methods

### Data

We derive polygenic scores for 245 phenotypes using the UK Biobank (UKBB) data only (Bycroft *et al*. 2018). We read dosages data from UKBB BGEN files using function `snp_readBGEN()` of R package bigsnpr (Privé *et al.* 2018). We divide the UKBB data in eight ancestry groups (Supplementary Note), and restrict to 437,669 individuals without second-degree relatives (KING kinship *<* 2^*−*3.5^). We also define a ninth ancestry group composed of 1709 unrelated Ashkenazi (see Methods below). For the variants, we use 1,040,096 HapMap3 variants used in the LD reference provided in Privé *et al.* (2020b) and that were also present in the iPSYCH2015 data (Bybjerg-Grauholm *et al.* 2020) with imputation INFO score larger than 0.6. Even though the iPSYCH data is not used in this study, we plan to use the PGS derived here for iPSYCH in the future.

To define phenotypes, we first map ICD10 and ICD9 codes (UKBB fields 40001, 40002, 40006, 40013, 41202, 41270 and 41271) to phecodes using R package PheWAS (Carroll *et al.* 2014; Wu *et al.* 2019). We filter down to 142 phecodes of interest that showed potential genetic signals in the PheWeb results from the SAIGE UKBB GWAS (Zhou *et al.* 2018; Taliun *et al.* 2020). There are 106 phecodes with sufficient power for penalized regression to include at least a few variants in the predictive models. We then look closely at all 2408 UKBB fields that we have access to and filter down to defining 111 continuous and 28 binary phenotypes based on manual curation. Description of the 245 phenotypes used in this study can be downloaded at https://github.com/privefl/UKBB-PGS/blob/main/phenotype-description.xlsx.

### Additional data: genotyped data

For the genotyped data used in some follow-up analyses, we restrict to variants that have been genotyped on both chips used by the UK Biobank, that pass quality control (QC) for all batches (cf. https://biobank.ctsu.ox.ac.uk/crystal/crystal/auxdata/ukb_snp_qc.txt) and QC for possible mismappings (Kunert-Graf *et al.* 2020), with a minor allele frequency (MAF) larger than 0.01 and imputation INFO score of 1. There are 586,534 such high-quality variants, which we read from the BGEN imputed data so that there is no missing value.

### Additional data: 8M+ variants

We also design a larger set of imputed variants to compare against using only HapMap3 variants for prediction. We first restrict to UKBB variants with MAF > 0.01 and INFO > 0.6. We then compile frequencies and imputation INFO scores from other datasets, iPSYCH and summary statistics for breast cancer, prostate cancer, coronary artery disease and type-1 diabetes (Bybjerg-Grauholm *et al.* 2020; Michailidou *et al.* 2017; Schumacher *et al.* 2018; Nikpay *et al.* 2015; Censin *et al.* 2017). We restrict to variants with a mean INFO > 0.5 in these other datasets, and also compute the median frequency. To exclude potential mismappings in the genotyped data (Kunert-Graf *et al.* 2020) that might have propagated to the imputed data, we compare median frequencies in the external data to the ones in UKBB (Figure S18). As we expect these potential errors to be localized around errors in the genotype data (confirmed in figure S19), we apply a moving-average smoothing on the frequency differences to increase power to detect these errors and also reduce false positives. We define the threshold on these smoothed differences based on visual inspection of their histogram. This is the same method we have previously applied to PC loadings to detect long-range LD regions when computing PCA (Privé *et al.* 2018, 2020a). This results in a set of 8,238,692 variants.

### Ashkenazi Jewish ancestry group

First, we refer the reader to the Supplementary Note on ancestry grouping for the details on how we define the other eight ancestry groups, and also to better understand how we infer the “Ashkenazi Jewish” ancestry group. Briefly, we project the UKBB data onto the PCA space of a reference dataset composed of many Jewish and non-Jewish individuals (Behar *et al*. 2013). We then compute the robust center (geometric median) of the Ashkenazi Jewish reference individuals, and compute the PC distance to this center for all projected UKBB individuals. Based on visual inspection of the histogram of these distances and on the fact that the closest non-Ashkenazi Jewish reference individual, an Italian Jew (Figure S20), is at distance 12.7, we use a threshold of 12.5 under which to assign to the “Ashkenazi Jewish” ancestry group. 1709 unrelated UKBB individuals are then assigned to this group. Note that, within the already defined eight ancestry groups, the closest individual to this new group belongs to the Italian group, and is at distance 17.3, therefore this new Ashkenazi group is not overlapping with any of the other groups defined previously.

### Penalized regression

To derive polygenic scores based on individual-level data from the UKBB, we use the fast implementation of penalized linear and logistic regressions from R package bigstatsr (Privé *et al.* 2019). We have also considered the recently developed R package snpnet for fitting penalized regressions on large genetic data; however, we provide theoretical and empirical evidence that bigstatsr is much faster than snpnet (Supplementary Note). Our implementation allows for lasso and elastic-net penalizations; yet, for the sake of simplicity and because the UKBB data is very large, we have decided to only use the lasso penalty (Privé *et al.* 2019). We recall that fitting a penalized linear regression with lasso penalty corresponds to finding the vector of effects *β* (also *µ* and *γ*) that minimizes

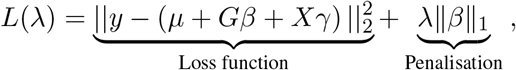

where *µ* is an intercept, *G* is the genotype matrix, *X* is the matrix of covariates, *y* is the (quantitative) phenotype of interest and *λ* is a hyper-parameter that controls the strength of the regularization and needs to be chosen. We use sex (Field 22001), age (Field 21022), birth date (Fields 34 & 52), Townsend deprivation index (Field 189) and the first 16 genetic principal components (Field 22009, Privé *et al*. (2020a)) as unpenalized covariates when fitting the lasso models.

We have extended our implementation in two ways by allowing for using different penalties for the variants (i.e. having _*j*_ *λ*_*j*_|*β*_*j*_| instead of *λ β* _1_). First, this enables us to use a different scaling for genotypes. By default, variants in *G* are implicitly scaled. By using *λ*_*j*_ *∝* (SD_*j*_)^(*ξ−*1)^, this effectively scales variant *j* by dividing it by (SD_*j*_)^*ξ*^ in our implementation. The default uses *ξ* = 1 but we also test *ξ* = 0 (no scaling) and *ξ* = 0.5 (Pareto scaling). We introduce a new parameter `power_scale` for which the user can provide a vector of values to test; the best value is chosen within the Cross-Model Selection and Averaging (CMSA) procedure (Privé *et al.* 2019). We also introduce a second parameter, `power_adaptive`, which can be used to put less penalizition on variants with the largest marginal effects (Zou 2006); we try 3 values here (0 the default, 0.5 and 1.5) and the best one is also chosen within the CMSA procedure.

### LDpred2-auto

Using the individual-level data from the training set in the UK biobank, we run a linear regression GWAS using function `big_univLinReg` of R package bigstatsr (Privé *et al.* 2018), accounting for the same covariates as in the penalized regression above. As LD reference, we use the one provided in Privé *et al*. (2020b) based on UKBB data for European ancestry. We use these summary statistics and this LD reference as input for LDpred2-auto. LDpred2 assumes a point-normal mixture distribution for effect sizes, where only a proportion of causal variants *p* contributes to the SNP heritability *h*^2^. In LDpred2-auto, these two parameters are directly estimated from the data (Privé *et al.* 2020b). We use the `sparse` option in LDpred2-auto to also obtain a vector of effects that is potentially sparse, i.e. effects of some variants are exactly 0. Also note that, as we use linear regression for all phenotypes, we use the total sample size instead of the effective sample size (4*/* (1*/n*_case_ + 1*/n*_control_)) for binary phenotypes as input to LDpred2. This means that heritability estimates from both LD score regression and LDpred2-auto must be transformed to the liability scale using both the prevalence in the GWAS and in the population; this can be performed using function `coef_to_liab` from R package bigsnpr. For simplicity, we assume here that the prevalence in the population is the same as the prevalence in the training set.

### New formula used in LDpred2

We also slightly modify the formula used in Privé *et al.* (2020b); we have previously used

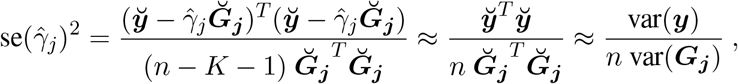

where 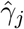 is the marginal effect of variant *j*, and where 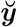 and **Ğ**_***j***_ are the vectors of phenotypes and genotypes for variant *j* residualized from *K* covariates, e.g. centering them. The first approximation expects 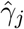 to be small, while the second approximation assumes the effects from covariates are small. However, we have found here that some variants can have very large effects, e.g. one variant explains about 30% of the variance in bilirubin log-concentration. Then, instead we compute

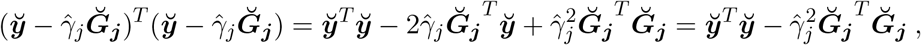

which now gives

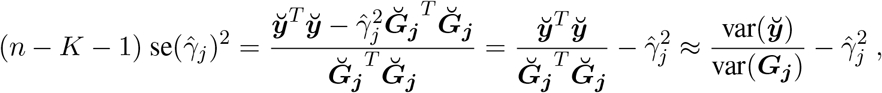

finally giving (note the added term 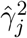)

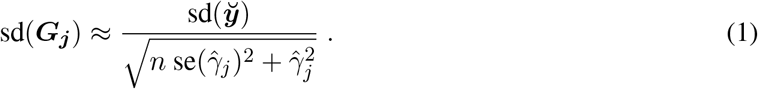

Figure S21 shows that the updated formula (1) is better; we now use it in the code of LDpred2, and also recommend using it for the QC procedure proposed in Privé *et al.* (2020b).

### Using more than HapMap3 variants in LDpred2

Here we also run LDpred2 using more than HapMap3 variants, based on a set of 8M+ variants (see above). However, LDpred2 cannot be run on 8M variants because the implementation is quadratic with the number of variants in terms of time and memory requirements. Thus, we employ another strategy consisting in keeping only the 1M most significant variants. To correct for winner’s curse, we employ the maximum likelihood estimator used in Zhong and Prentice (2008) and Shi *et al.* (2016):

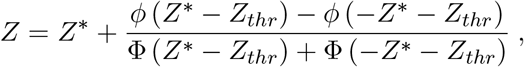

where *ϕ* is the standard normal density function, Φ is the standard normal cumulative density function, *Z* is the Z-score obtained from the GWAS, *Z*_*thr*_ is the threshold used on (absolute) Z-scores for filtering, and *Z*^*\**^ is the corrected Z-score that we estimate and use. As input for LDpred2, instead of using *β* (along with SE(*β*) and *N*), we use *β*^*\**^ = *β · Z*^*\**^*/Z* where *Z* = *β/*SE(*β*). This is now implemented in function `snp_thr_correct` of package bigsnpr.

### Performance metric

Here we use the partial correlation as the performance metric, which is the correlation between the PGS and the phenotype after they have been both residualized using the covariates used in this paper, i.e. sex, age, birth date, deprivation index and 16 PCs. To derive 95% confidence intervals for these correlations, we use Fisher’s Z-transformation. We implement this in function `pcor` of R package bigstatsr and use it here.

## Supporting information

Supplementary Note on ancestry grouping

Supplementary Note on snpnet comparison

## Data Availability

The UK Biobank resource is available to bona fide researchers for health-related research in the public interest. All researchers who wish to access the research resource must register with UK Biobank by completing the registration form at https://www.ukbiobank.ac.uk/enable-your-research/register.

## Code and results availability

All code used for this paper is available at https://github.com/privefl/UKBB-PGS/tree/master/code. Links to the code used for the two supplementary notes are provided there. We have extensively used R packages bigstatsr and bigsnpr (Privé *et al*. 2018) for analyzing large genetic data, packages from the future framework (Bengtsson 2020) for easy scheduling and parallelization of analyses on the HPC cluster, and packages from the tidyverse suite (Wickham *et al.* 2019) for shaping and visualizing results. We have also used R package deming for fitting Deming regressions.

R packages bigstatsr and bigsnpr can be installed from GitHub and CRAN. A tutorial on fitting penalized regressions with R package bigstatsr is available at https://privefl.github.io/bigstatsr/articles/penalized-regressions.html. A tutorial on running LDpred2 with R package bigsnpr is available at https://privefl.github.io/bigsnpr/articles/LDpred2.html.

PC centers of the individuals from the nine ancestry groups derived here can be downloaded at https://github.com/privefl/UKBB-PGS/blob/main/pop_centers.csv. Description of the 245 phenotypes used in this study can be downloaded at https://github.com/privefl/UKBB-PGS/blob/main/phenotype-description.xlsx. Effect sizes for 215 polygenic scores derived in this study can be downloaded at https://figshare.com/articles/dataset/Effect_sizes_for_215_polygenic_scores/14074760.

## Acknowledgements

Authors thank Abdel Abdellaoui for his help with defining the “years of education” phenotype, and Alex Diaz-Papkovich and others for their useful feedback on the ancestry inference. Authors thank GenomeDK and Aarhus University for providing computational resources and support that contributed to these research results. This research has been conducted using the UK Biobank Resource under Application Number 58024.

