## Supplementary Materials for "High-resolution portability of 245 polygenic scores when derived and applied in the same cohort"

### Supplementary Tables and Figures

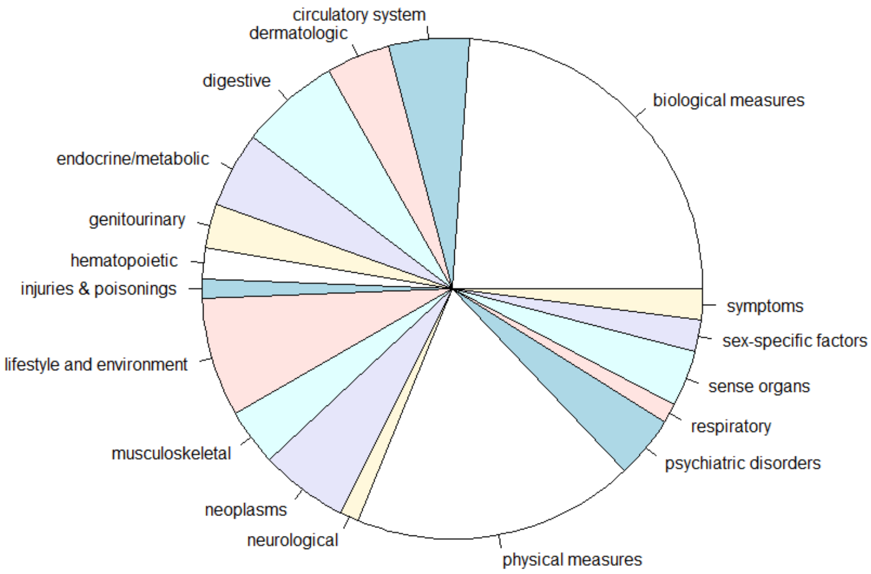


Figure S1: Pie chart of the categories of the 245 phenotypes used in this study. A full description of these phenotypes can be downloaded at `https://github.com/privefl/UKBB-PGS/blob/main/phenotype-description.xlsx`.

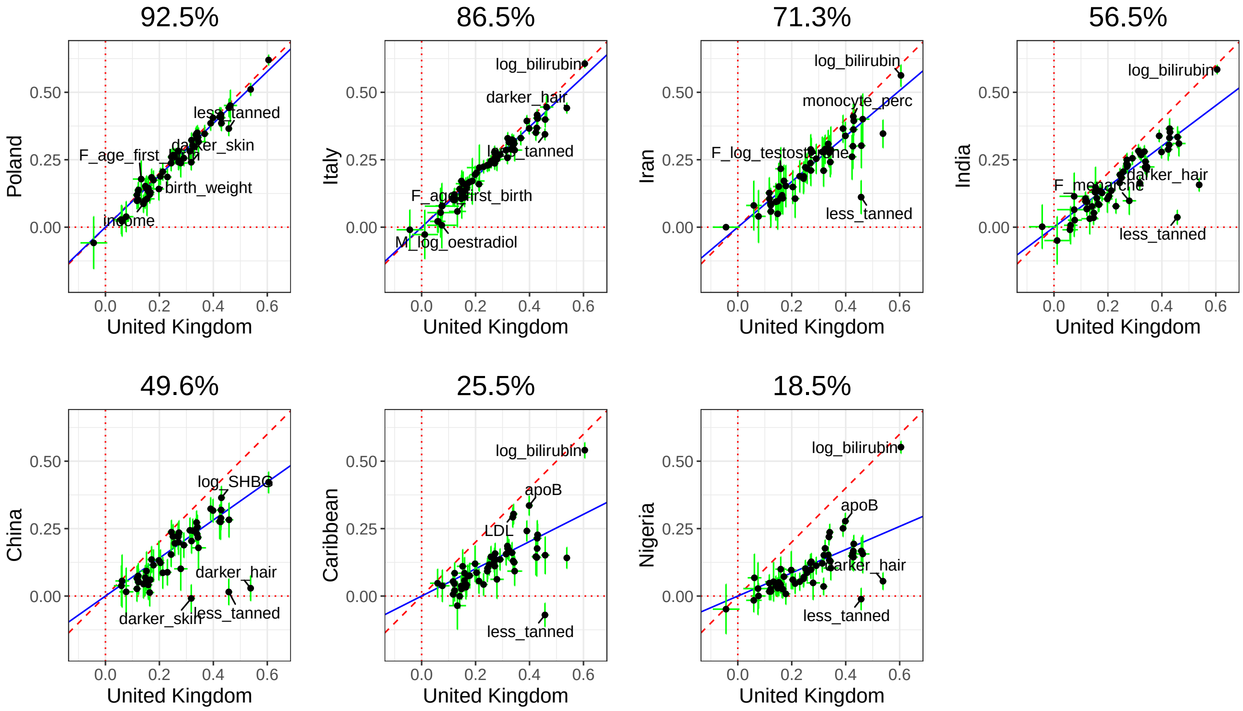


Figure S2: Partial correlation (and 95% CI) in the UK test set versus in a test set from another ancestry group. Each point represents a phenotype (only 83 of the continuous phenotypes here) and training has been performed with penalized regression on UK individuals (training 1 in table 1) and **genotyped** variants. The slope (in blue) is computed using Deming regression accounting for standard errors in both x and y, fixing the intercept at 0. The square of this slope is provided above each plot, which we report as the relative predictive performance compared to testing in UK.

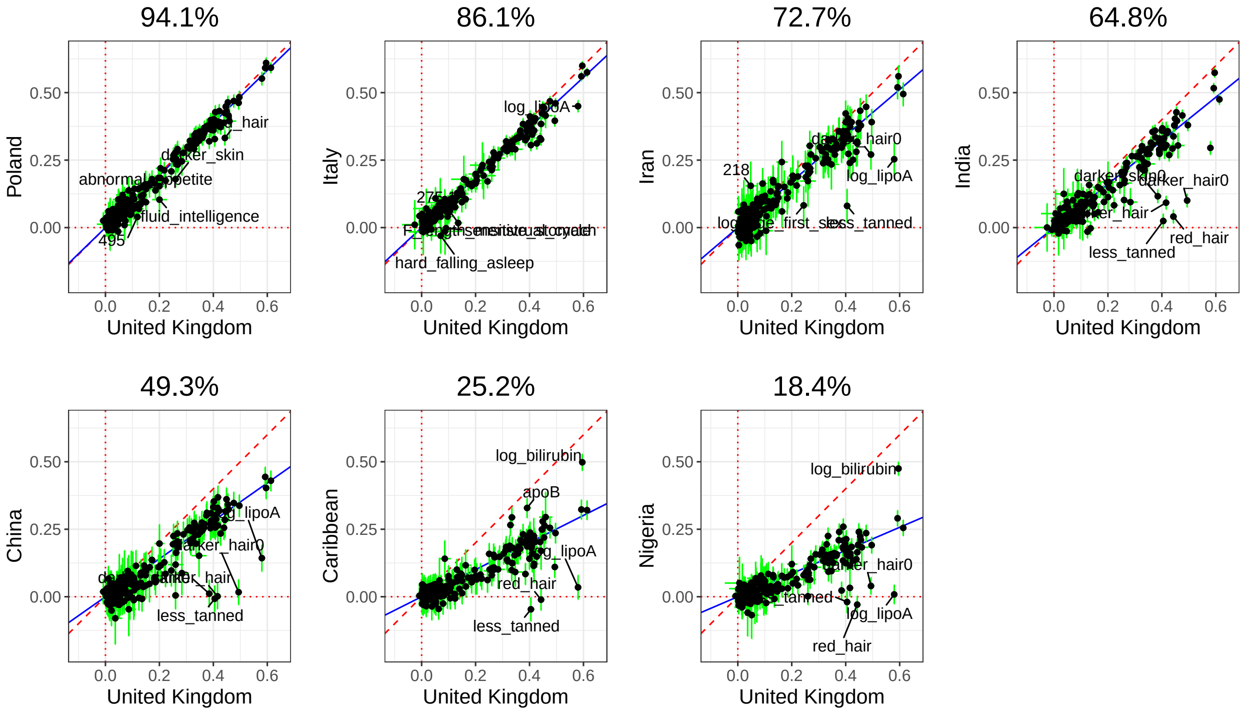


Figure S3: Partial correlation (and 95% CI) in the UK test set versus in a test set from another ancestry group. Each point represents a phenotype and training has been performed with **LDpred2-auto** on UK individuals (training 1 in table 1) and HapMap3 variants. The slope (in blue) is computed using Deming regression accounting for standard errors in both x and y, fixing the intercept at 0. The square of this slope is provided above each plot, which we report as the relative predictive performance compared to testing in UK.

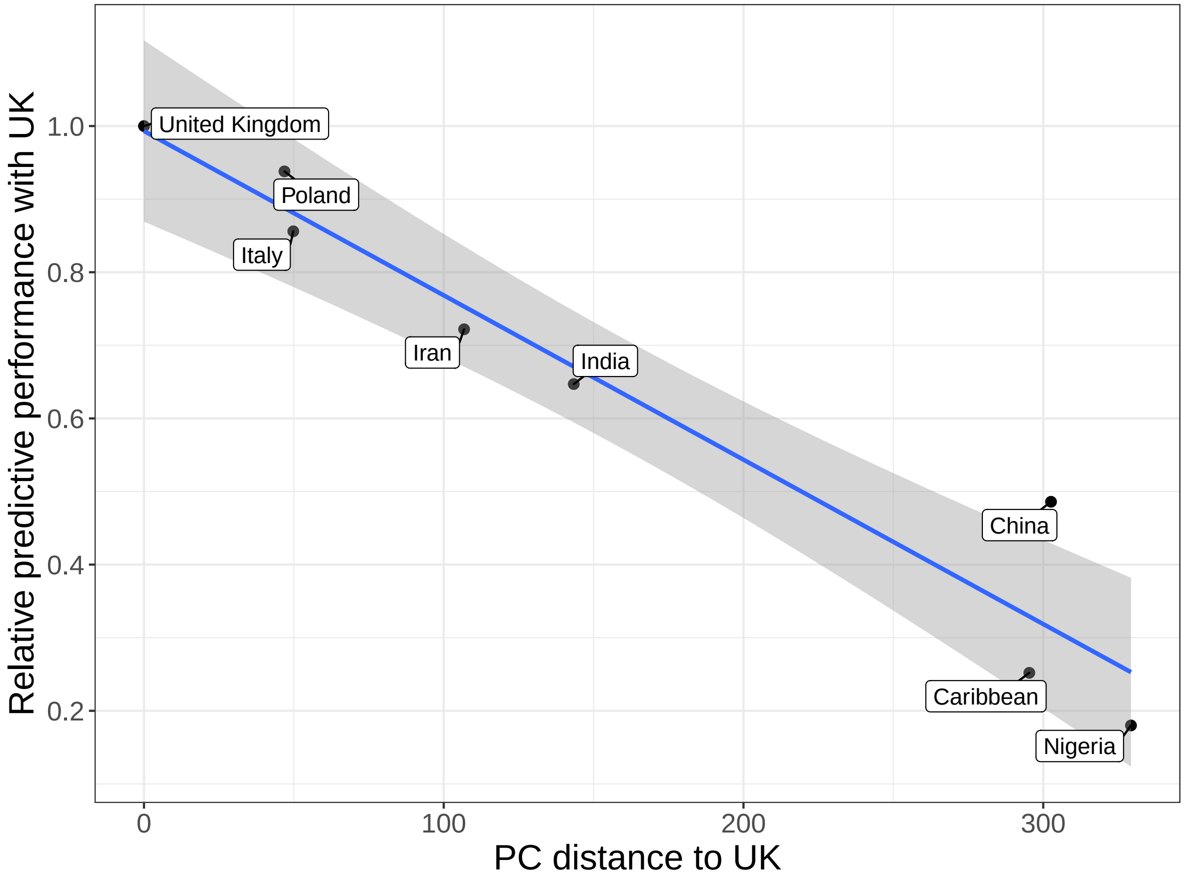


Figure S4: Relative predictive performance compared to the UK (ratio of variance explained in one group compared to in the UK group) versus PC distance from the UK. PCA is computed using individuals from test 1 (Table 1), and PC distances are computed using Euclidean distance between geometric medians of the first 32 PC scores of each ancestry group (shown in figure S5). Relative performance values are the ones reported in figure 2 of the main text. The slope and standard errors are computed internally by function `geom_smooth(method = "lm")` of R package ggplot2.

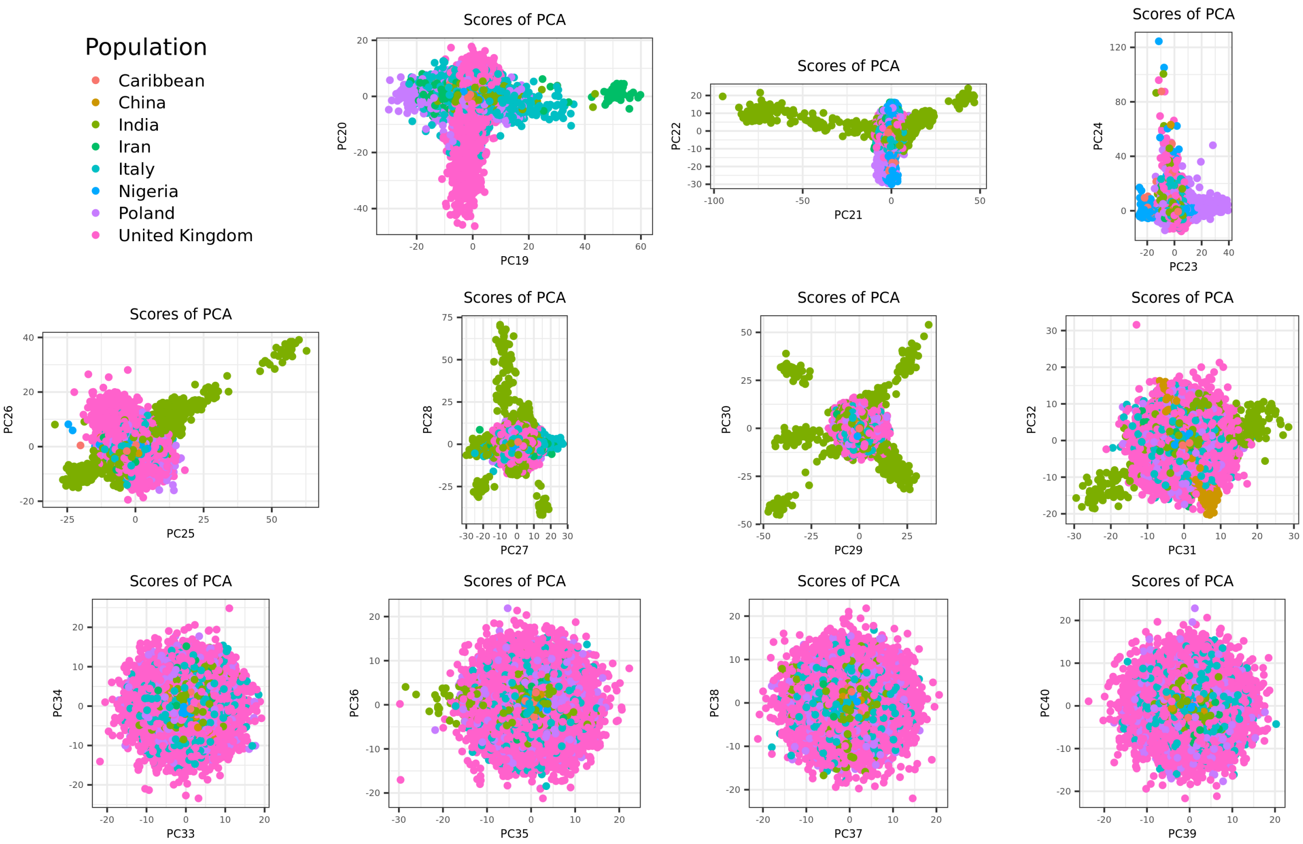


Figure S5: PC scores 19 to 40 when PCA is computed using individuals from test 1 (Table 1). PCs 19 to 32 visually capture some population structure, so we use first 32 PCs when computing the PC distances.

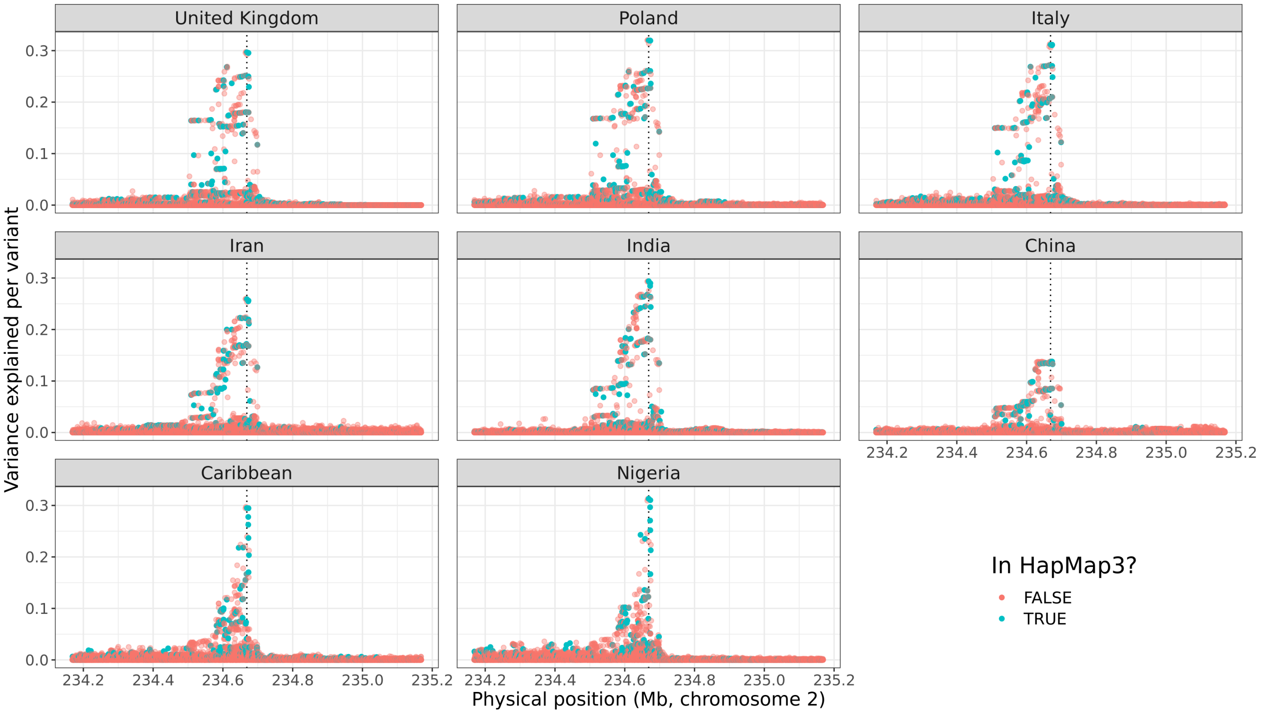


Figure S6: Zoomed Manhattan plot for total **bilirubin** concentration. The phenotypic variance explained per variant is computed as $r^2 = t^2/(n+t^2)$, where $t$ is the t-score from GWAS and $n$ is the degrees of freedom (the sample size minus the number of variables in the model, i.e. the covariates used in the GWAS, the intercept and the variant). The GWAS includes all variants with an imputation INFO score larger than 0.3 and within a 500Kb radius around the top hit from the GWAS performed in the UK training set and on the HapMap3 variants, represented by a vertical dotted line.

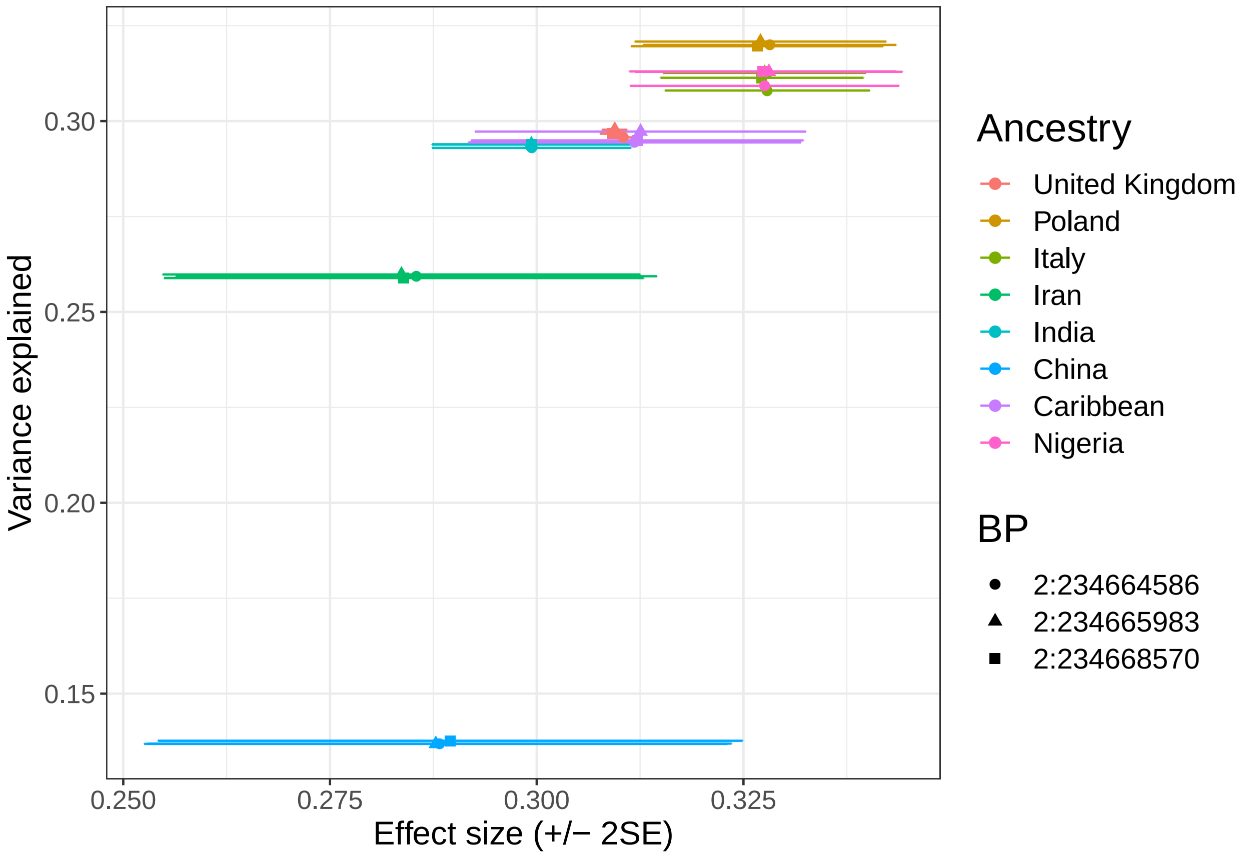


Figure S7: Effect sizes and variance explained for the top three variants from figure S6.

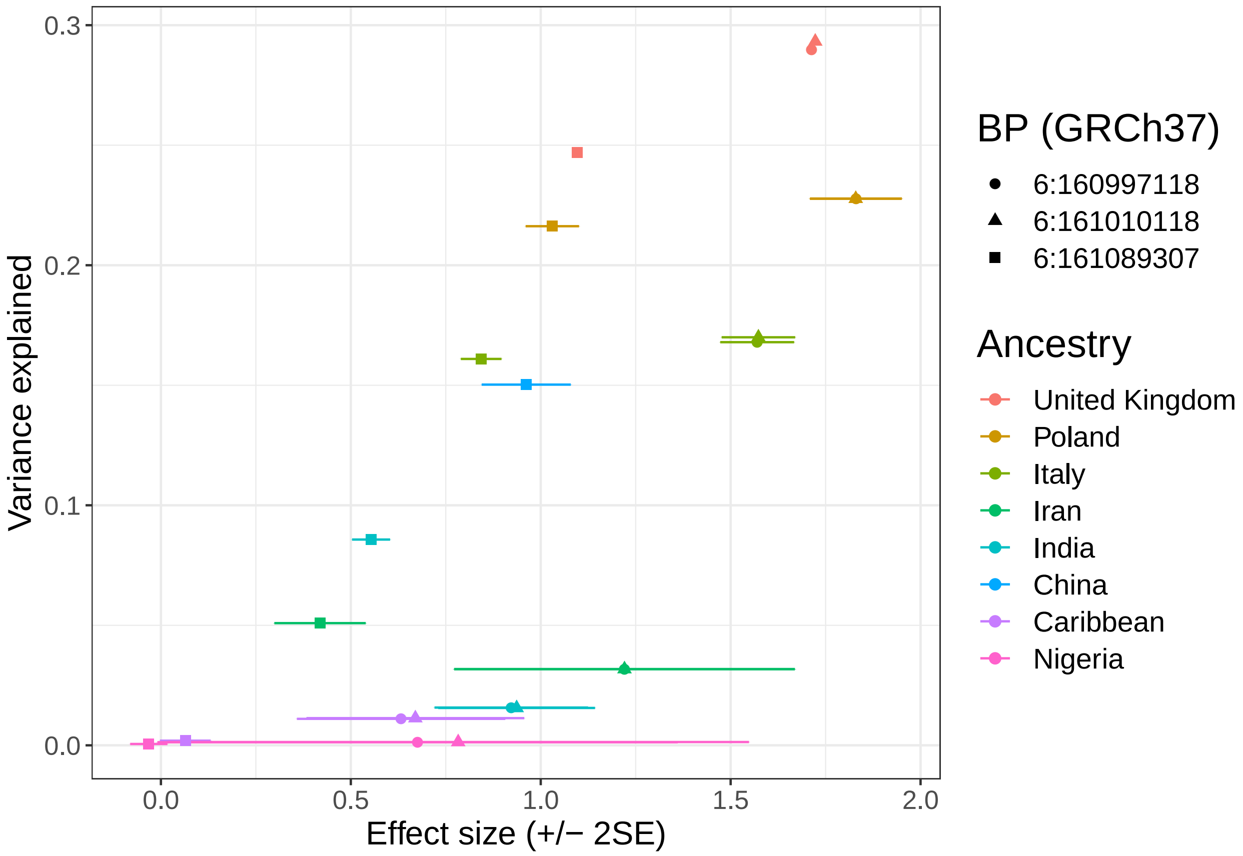


Figure S8: Effect sizes and variance explained for the top three variants from figure 4.

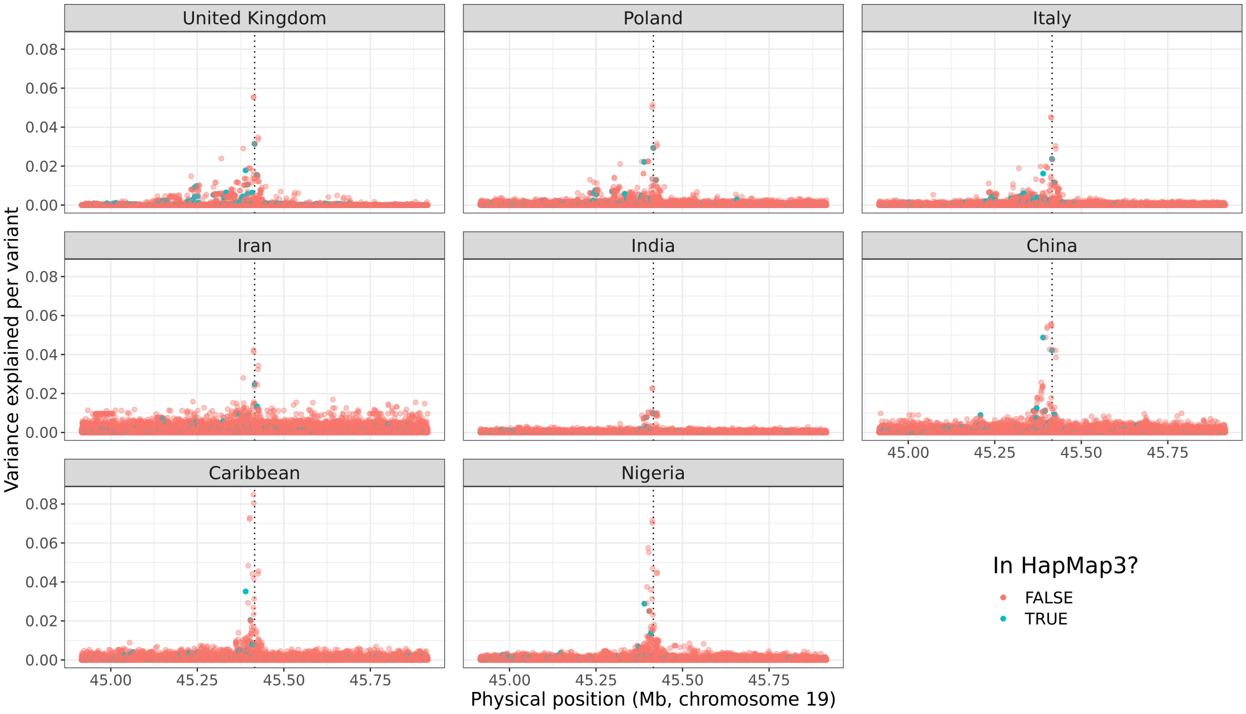


Figure S9: Zoomed Manhattan plot for **apolipoprotein B** concentration. The phenotypic variance explained per variant is computed as $r^2 = t^2/(n + t^2)$, where $t$ is the t-score from GWAS and $n$ is the degrees of freedom (the sample size minus the number of variables in the model, i.e. the covariates used in the GWAS, the intercept and the variant). The GWAS includes all variants with an imputation INFO score larger than 0.3 and within a 500Kb radius around the top hit from the GWAS performed in the UK training set and on the HapMap3 variants, represented by a vertical dotted line.

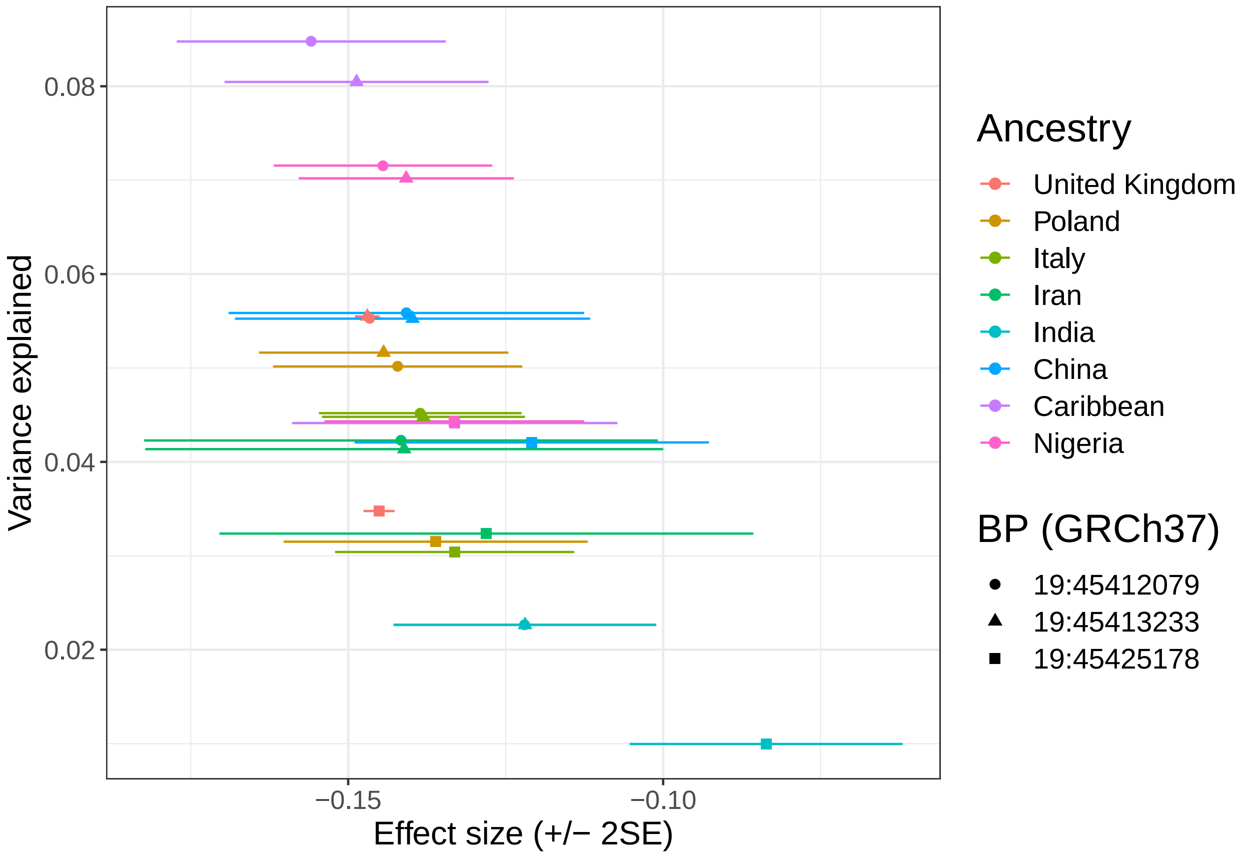


Figure S10: Effect sizes and variance explained for the top three variants from figure S9.

| Phenotype | Set of variants | h2 [2.5%-97.5%] | p [2.5%-97.5%] |
|---|---|---|---|
| 174.1 | top1M | 0.0889 [0.086-0.092] | 0.0076 [0.00678-0.00841] |
| 174.1 | HM3 | 0.0299 [0.0264-0.0334] | 0.000881 [0.000636-0.00117] |
| 185 | top1M | 0.113 [0.109-0.116] | 0.00819 [0.00743-0.00906] |
| 185 | HM3 | 0.0381 [0.0343-0.0423] | 0.000784 [0.000588-0.00105] |
| 411.4 | top1M | 0.0641 [0.0624-0.0659] | 0.0152 [0.0138-0.0168] |
| 411.4 | HM3 | 0.0401 [0.0379-0.0422] | 0.00457 [0.00397-0.00526] |
| apoB | top1M | 0.269 [0.265-0.272] | 0.0533 [0.0498-0.0568] |
| apoB | HM3 | 0.163 [0.16-0.166] | 0.00132 [0.00119-0.00145] |
| height | top1M | 0.482 [0.479-0.486] | 1 [1-1] |
| height | HM3 | 0.546 [0.541-0.552] | 0.0226 [0.0218-0.0235] |
| log_bilirubin | top1M | 0.301 [0.267-0.363] | 0.214 [0.195-0.227] |
| log_bilirubin | HM3 | 0.361 [0.357-0.365] | 0.000481 [0.000423-0.000545] |
| log_BMI | top1M | 0.173 [0.171-0.176] | 1 [1-1] |
| log_BMI | HM3 | 0.263 [0.26-0.266] | 0.0426 [0.0404-0.0446] |
| log_lipoA | top1M | 0.696 [0.689-0.702] | 0.0116 [0.011-0.0122] |
| log_lipoA | HM3 | 0.34 [0.336-0.345] | 0.000229 [0.000192-0.000268] |

Table S1: Estimates of SNP heritability $h^2$ and proportion of causal variants $p$ from LDpred2-auto, when using either 1,040,096 HapMap3 variants or when prioritizing 1M variants out of 8M+ common variants, for eight phenotypes. Quantiles of all estimates are also reported.

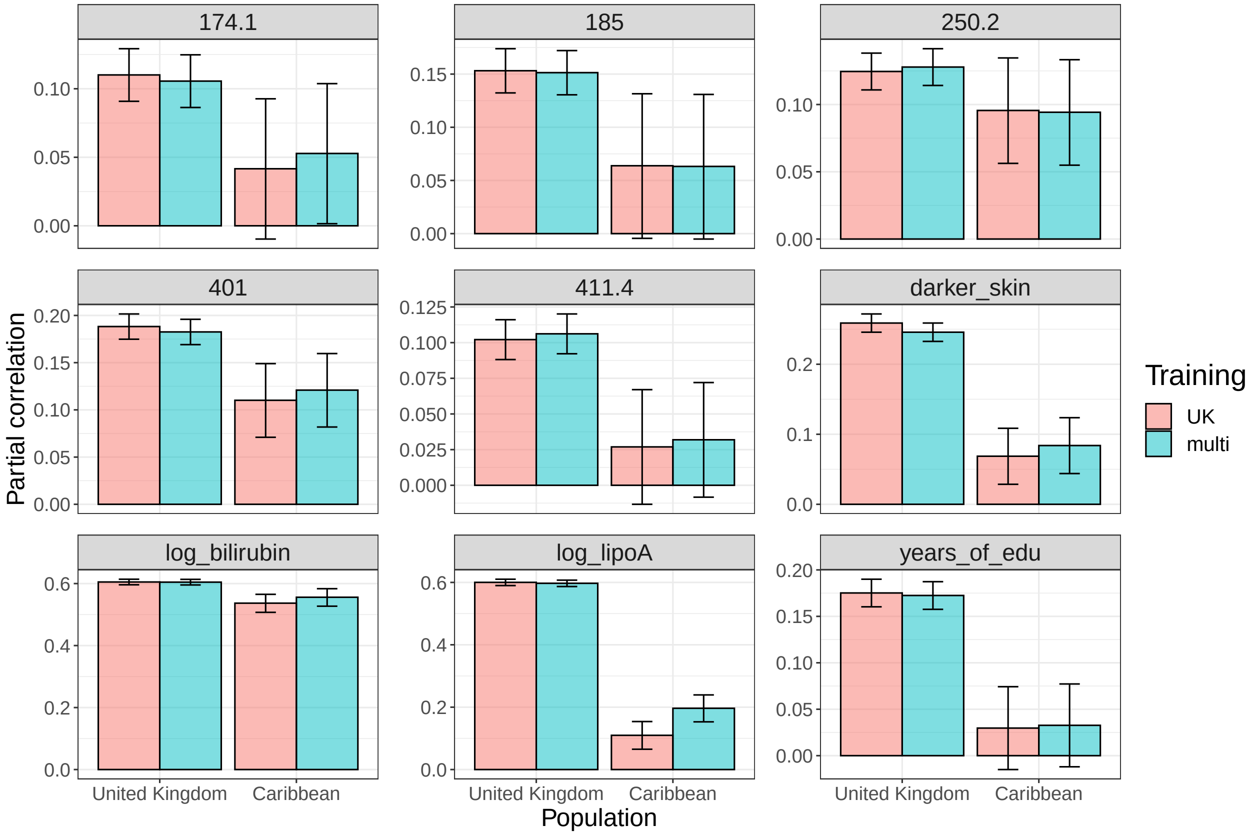


Figure S11: Partial correlation achieved per phenotype (each panel) and per ancestry group (x-axis) when training penalized regressions either with UK individuals only (training 1 in table 1) or when using individuals of multiple ancestries (training 2).

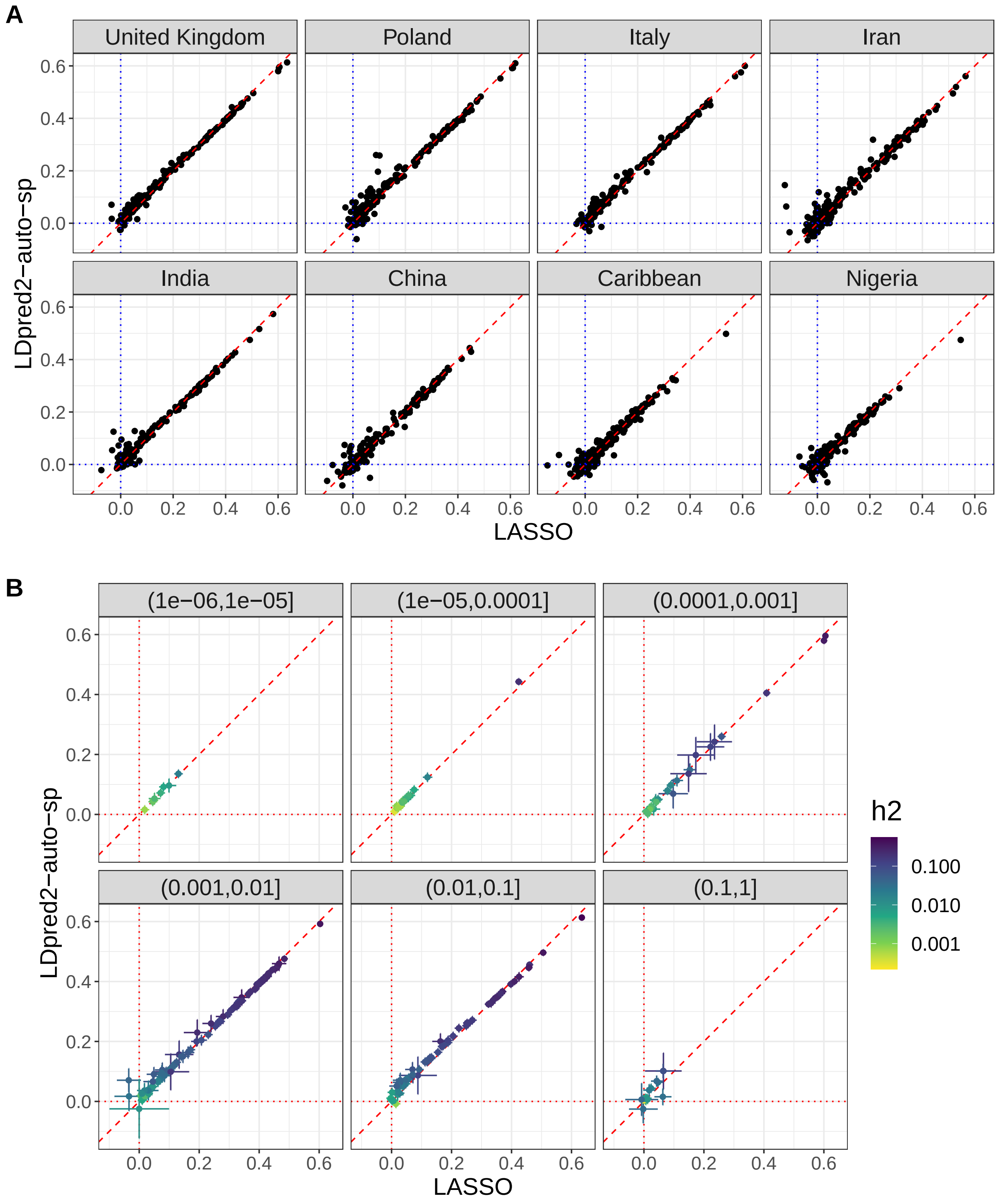


Figure S12: **A)** Partial correlations (and 95% CI) achieved per phenotype (each point) and per ancestry group (each panel) when training either with LASSO or with LDpred2-auto. **B)** Focusing now on the UK panel from A), each panel represents a range of proportion of causal variants $p$ and points are colored by SNP heritability $h^2$ (estimates from LDpred2-auto). Penalized regression tends to provide better predictive performance than LDpred2 for phenotypes for which partial-$r > 0.3$, and inversely.

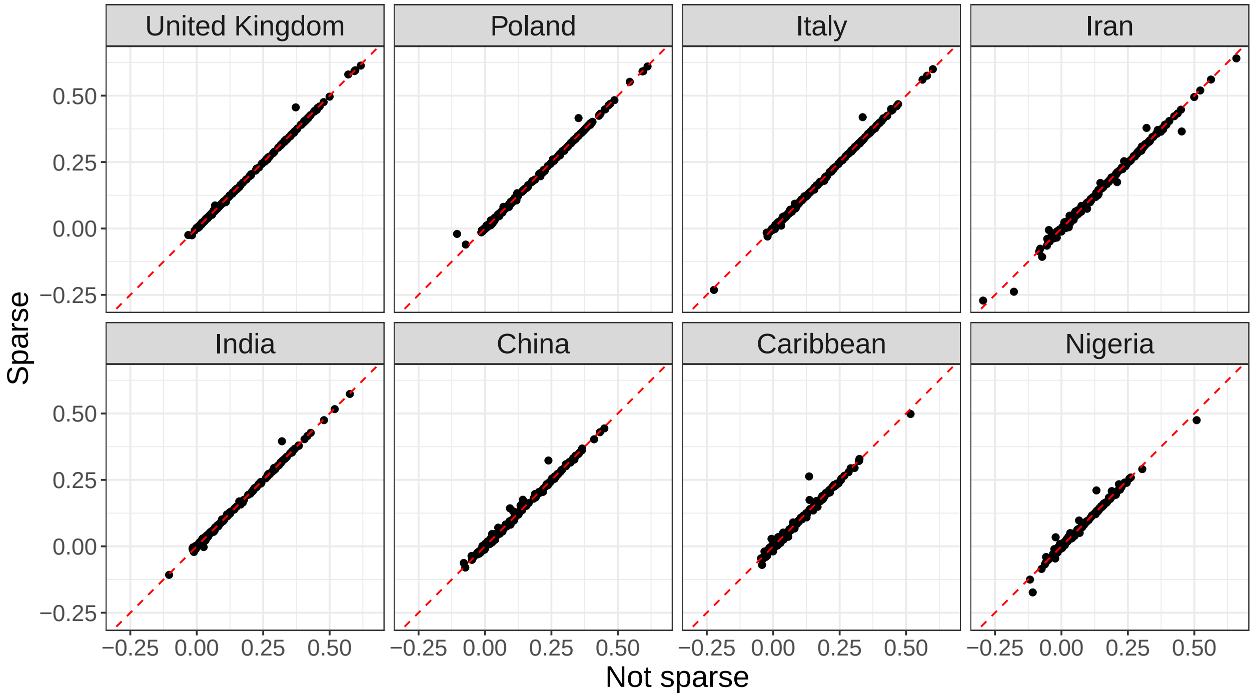


Figure S13: Partial correlations achieved per phenotype (each point) and per ancestry group (each panel) when training either with LDpred2-auto or with LDpred2-auto-sparse (sparse option enabled).

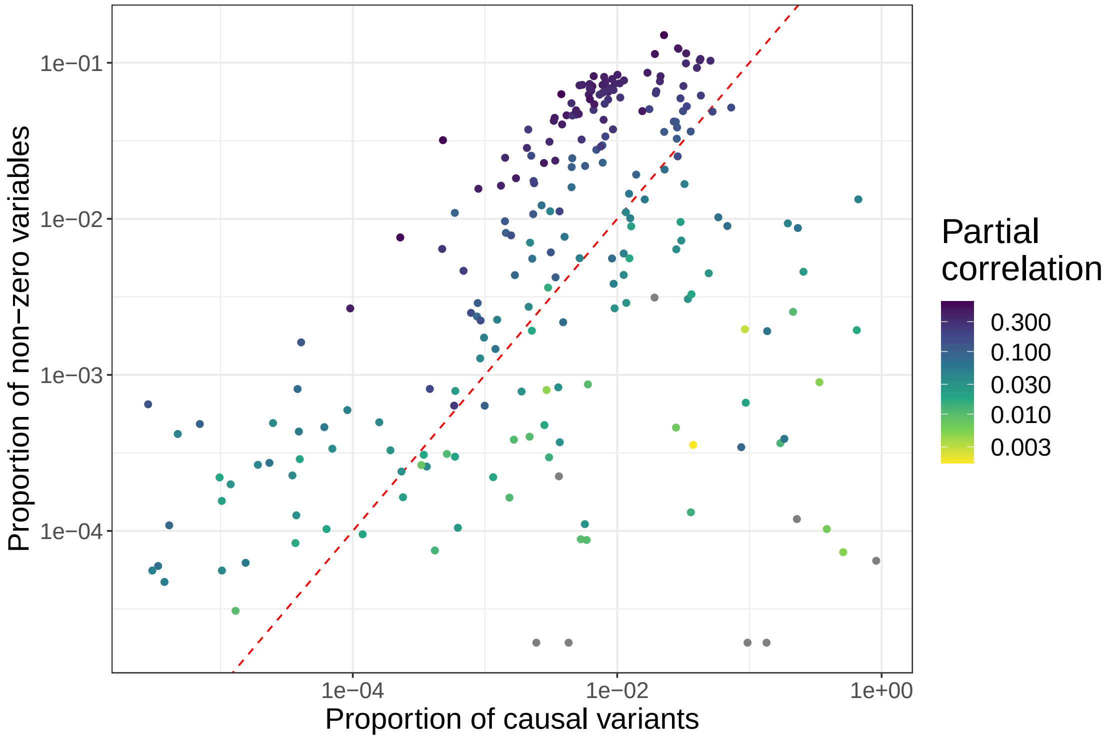


Figure S14: Proportion of variants with non-zero effects in the penalized regression models for each phenotype (point) versus the proportion of causal variants $p$ estimated from LDpred2-auto, colored by the partial correlation achieved in the UK test set.

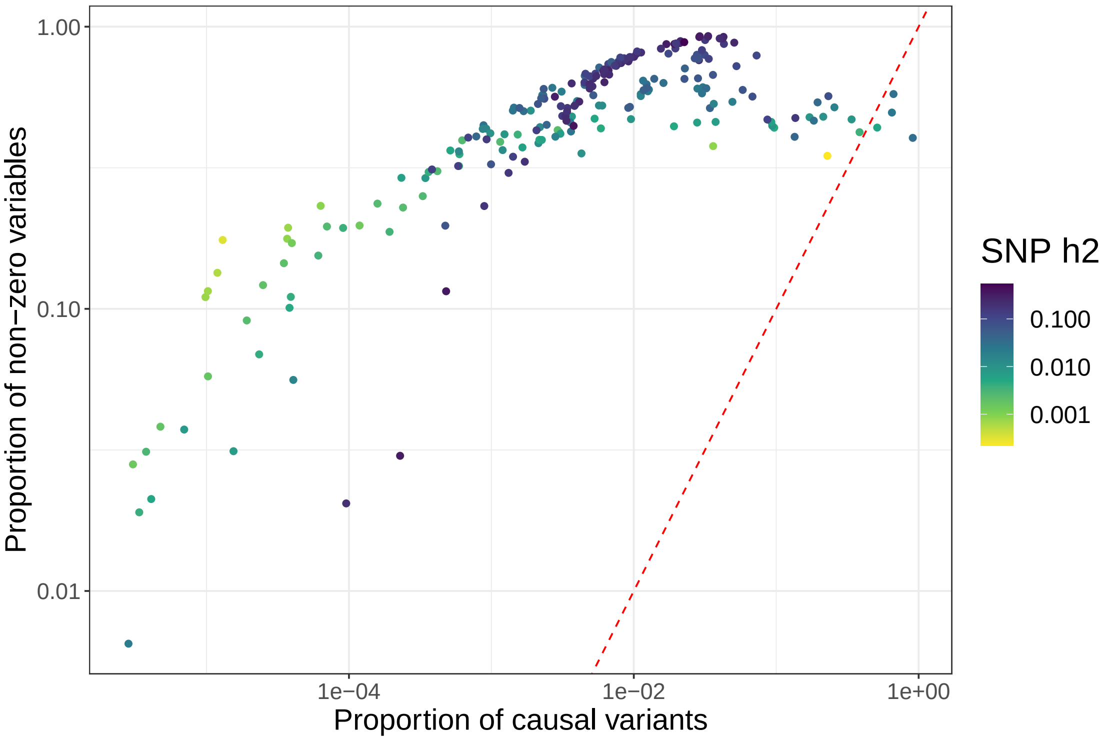


Figure S15: Proportion of variants with non-zero effects in LDpred2-auto-sparse for each phenotype (point) versus the proportion of causal variants $p$ estimated from LDpred2-auto, colored by the SNP heritability $h^2$ estimated from LDpred2-auto.

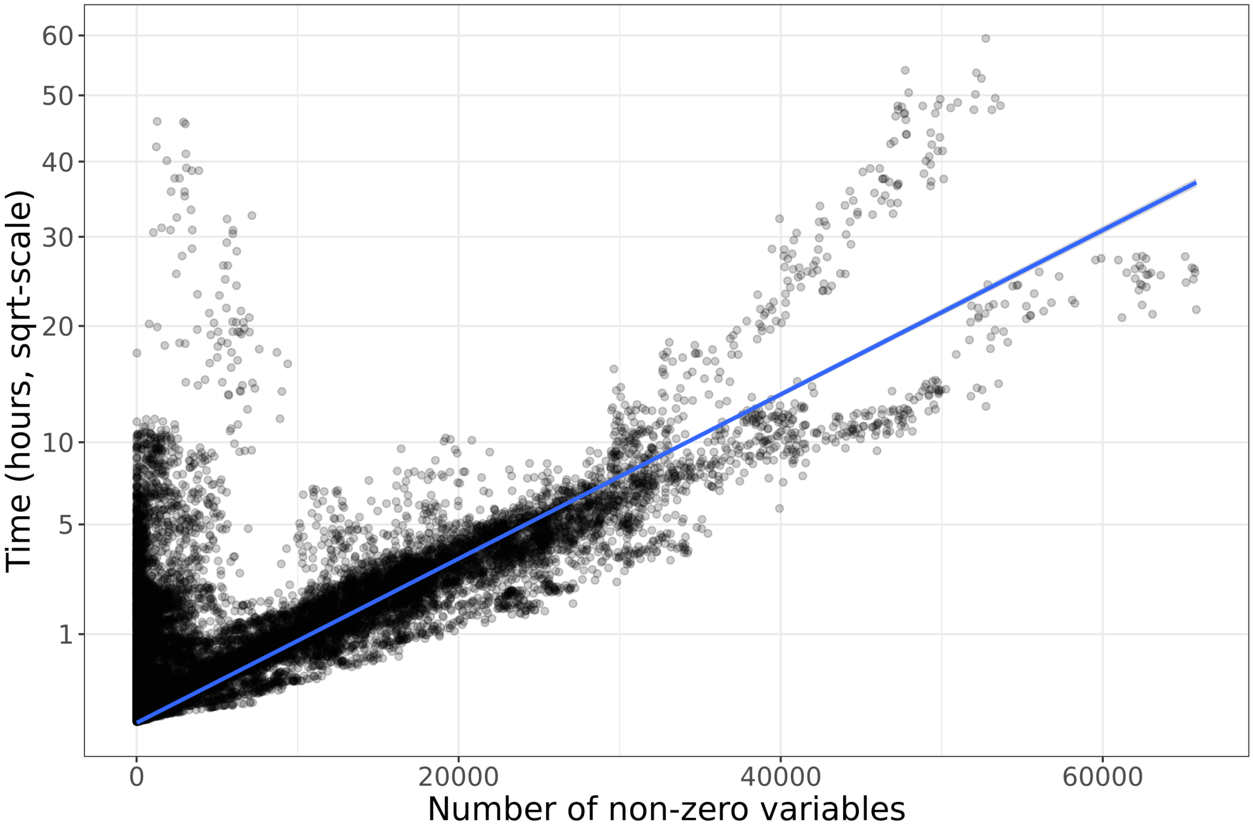


Figure S16: Computation times for all penalized regression models run using the 1M HapMap3 variants. We recall that we usually run 90 models for each phenotype because we use 9 sets of hyper-parameters and K=10 folds. Computation time is largely quadratic with the number of non-zero effects in the model. It is also dependent on the compute node and the loading of the HPC cluster at the time of running (Figure S17).

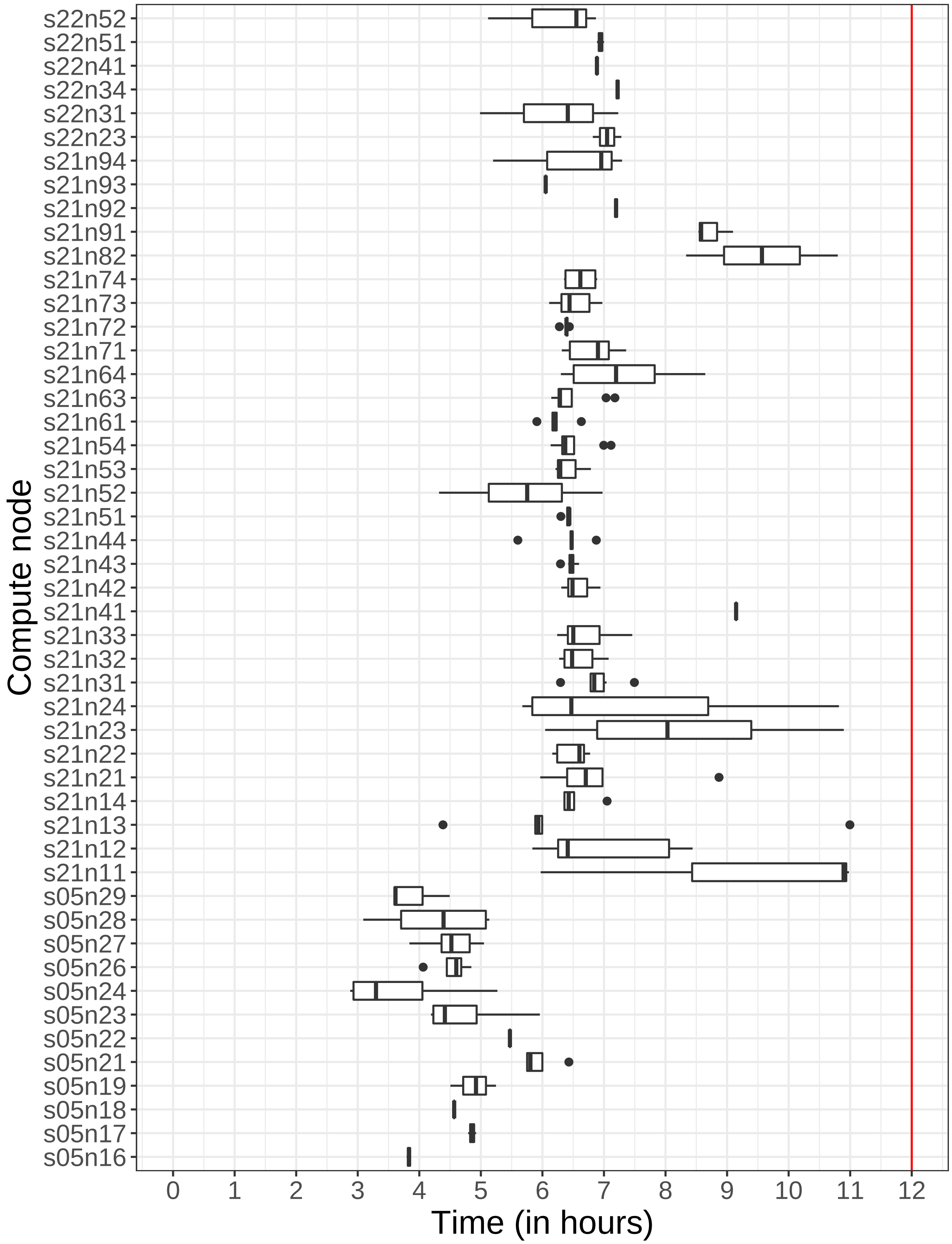


Figure S17: Computation times for fitting LDpred2-auto (with default 1000 burn-in iterations + 500 more + sparse option running 150 more) using the 1M HapMap3 variants. Running times should be the same for all phenotypes, yet we see some variability depending on the node used. Some fitting had to be run again because it exceeded the 12-hour timeout, which happened a few times when and the HPC cluster was particularly crowded.

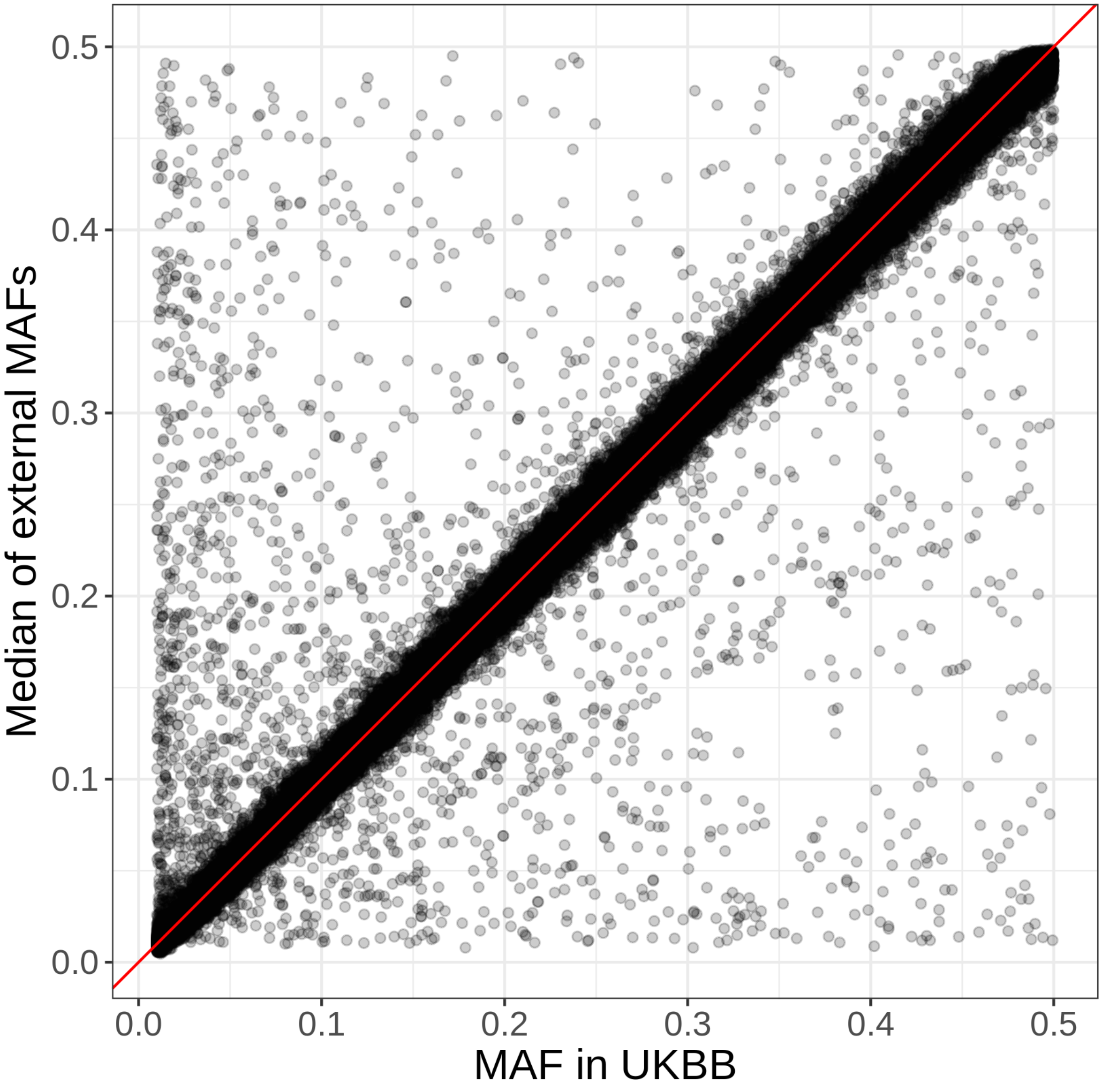


Figure S18: Comparison between frequencies in the UK Biobank and frequencies in external data.

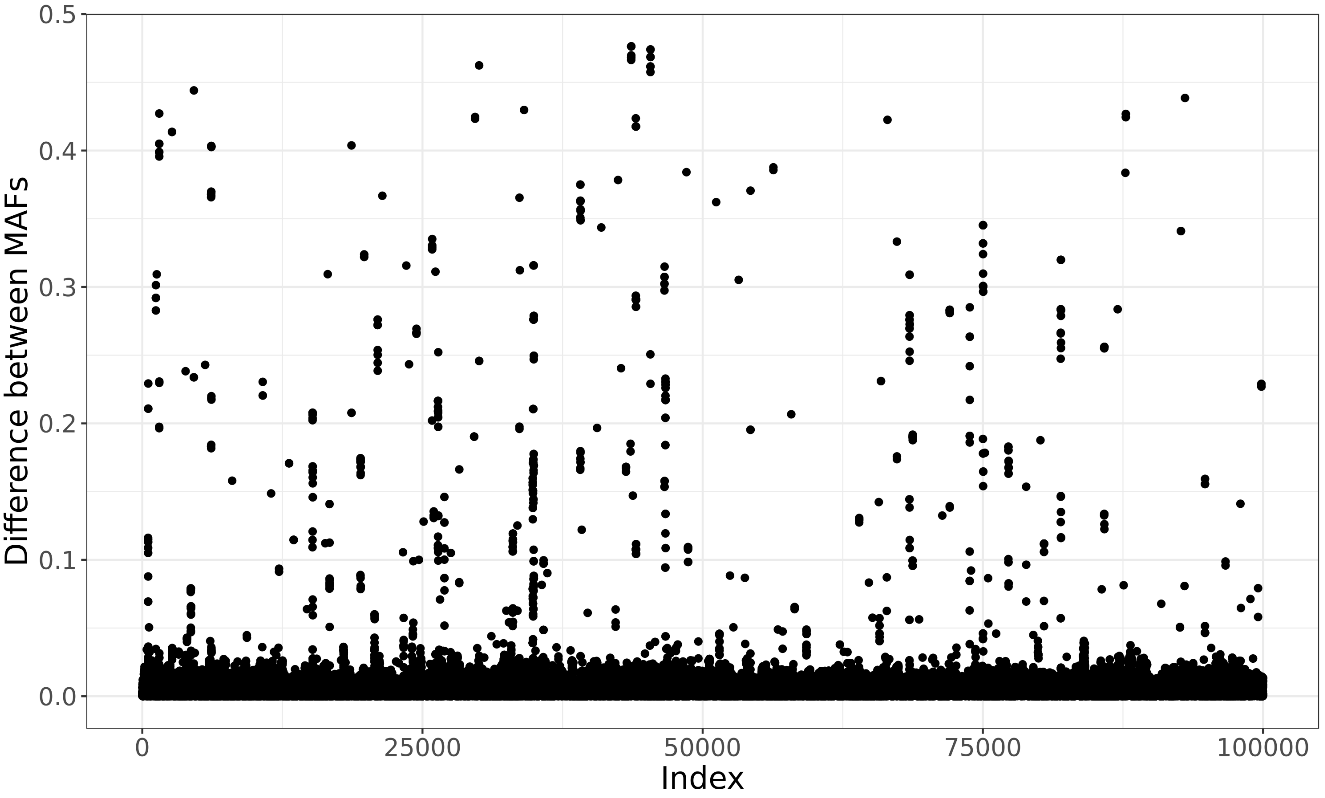


Figure S19: Differences in MAF between the first 100,000 variants in UK Biobank and external data. These differences (likely errors in UKBB) are hypothetically grouped around errors in the genotyped data that propagated to the imputed data.

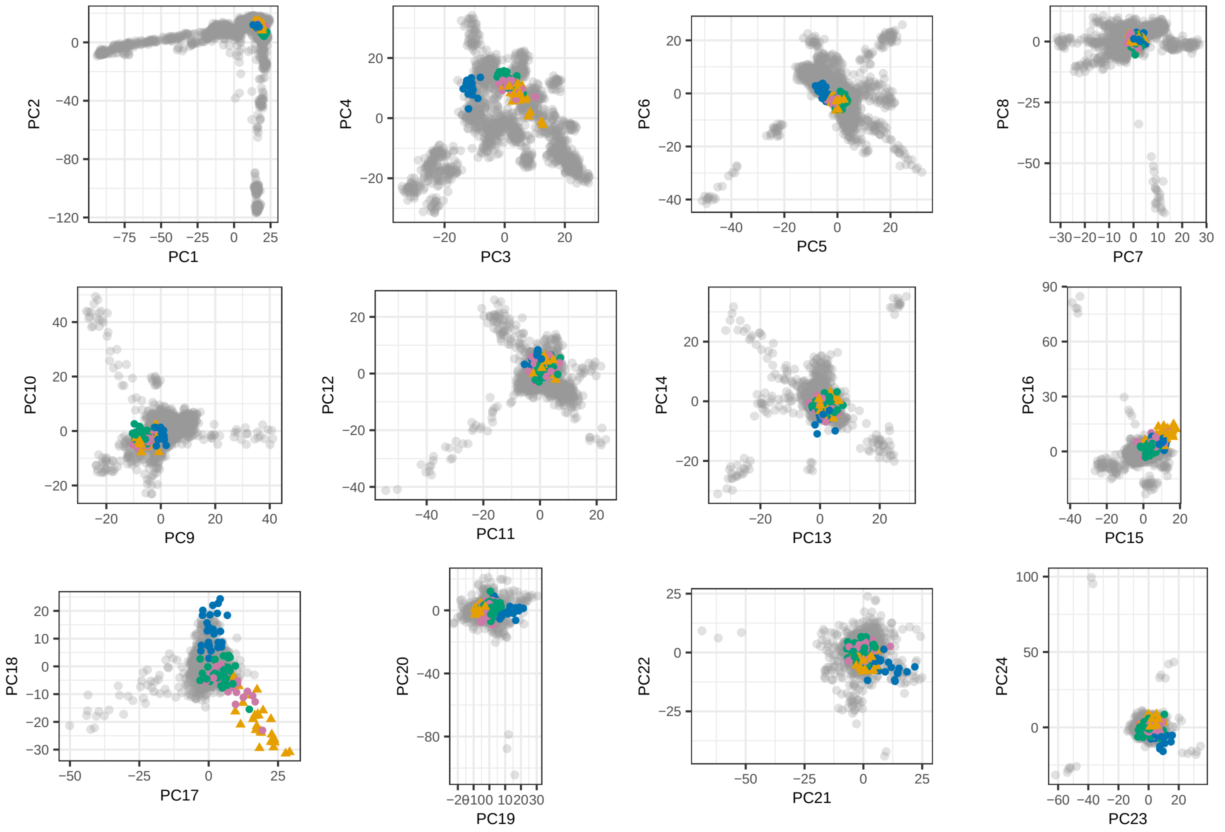


Figure S20: First 24 PC scores for the PCA computed in the reference dataset composed of several Jewish and non-Jewish individuals (Behar *et al.* 2013). Orange triangles represent the Ashkenazi Jews, pink points the Italian and Sephardi Jews, green points the Maghrebian Jews, and blue points the Iranian and Iraqi Jews.

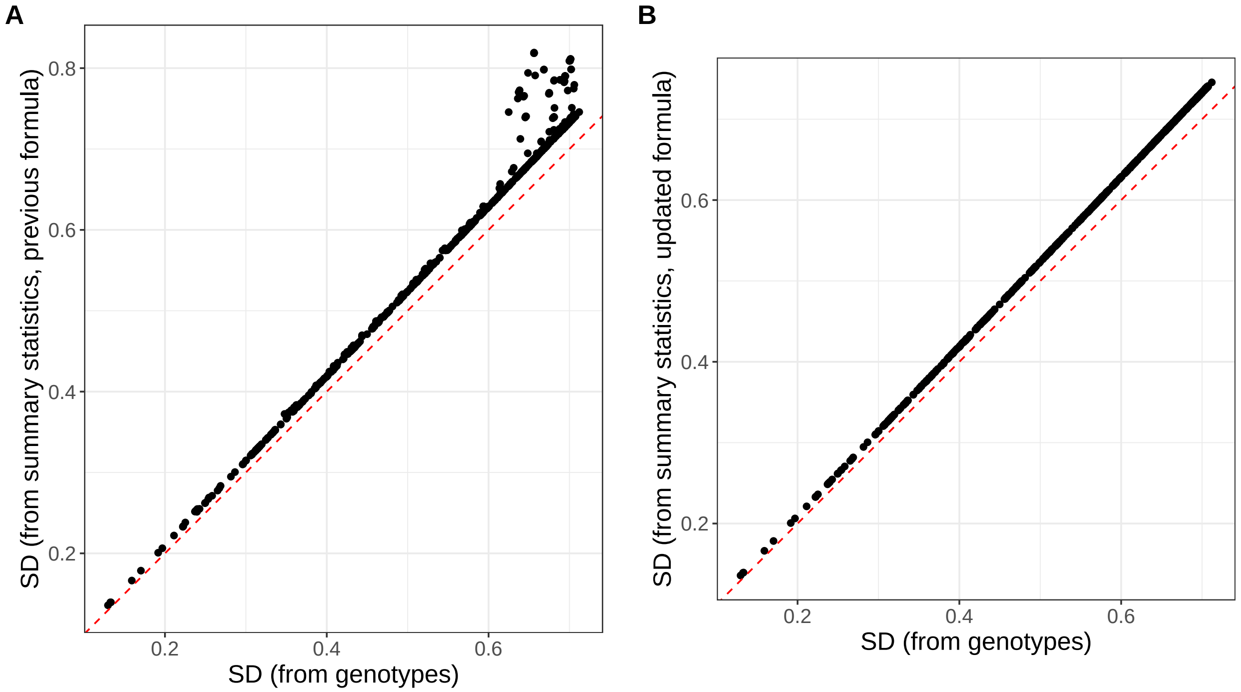


Figure S21: Comparison of the standard deviations (SD) computed from both genotypes and summary statistics for the 1000 most associated variants with bilirubin concentration. A) uses the previous formula $\text{sd}(\boldsymbol{G_j}) \approx \frac{\text{sd}(\boldsymbol{y})}{\sqrt{n\ \text{se}(\hat{\gamma}_j)^2}}$ proposed in Privé *et al.* (2020) while B) uses the updated formula $\text{sd}(\boldsymbol{G_j}) \approx \frac{\text{sd}(\boldsymbol{y})}{\sqrt{n\ \text{se}(\hat{\gamma}_j)^2 + \hat{\gamma}_j^2}}$ proposed here, which does one less approximation. The slope slightly larger than 1 can be explained by $\text{sd}(\boldsymbol{y}) > \text{sd}(\breve{\boldsymbol{y}})$.
